# Early onset leprosy reveals a joint effect of *LRRK2* and *NOD2* variants

**DOI:** 10.1101/2021.03.25.21253623

**Authors:** Monica Dallmann-Sauer, Yong Zhong Xu, Ana Lúcia França da Costa, Shao Tao, Wilian Correa-Macedo, Jérémy Manry, Laurent Abel, Alexandre Alcaïs, Aurélie Cobat, Vinicius M. Fava, Christian M. Probst, Marcelo T. Mira, Erwin Schurr

## Abstract

Leprosy, caused by *Mycobacterium leprae*, has a long incubation period and cases with age-of-onset <5 years are rare. Here, we studied a three-generational multiplex leprosy family which included monozygotic twins age <24 months suffering from paucibacillary leprosy. Whole genome sequencing identified a homozygous double mutation in the *LRRK2* gene (N551K, R1398H) and a heterozygous mutation in *NOD2* (R702W) as candidate variants underlying the early onset phenotype in the twins. The same amino acid substitutions had previously been identified as shared risk-modulating factors for Crohn’s disease and Parkinson’s disease. To evaluate the functional impact of the *LRRK2* mutations, we employed genome editing in RAW264.7 cells. Cells expressing the LRRK2 variants displayed reduced respiratory burst and apoptosis following mycobacterial challenge. Moreover, the BCG-induced respiratory burst was significantly lower in LRRK2 wild-type-expressing cells transfected with *NOD2* R702W compared with *NOD2* wild-type constructs. Employing co-immunoprecipitation, we showed that LRRK2 and NOD2 wild-type proteins interact in RAW cells. This interaction was independent of the LRRK2 variants but strongly reduced for NOD2 R702W. However, N-glycolyl MDP-triggered RIP2 phosphorylation and NF-kB activation were additively reduced by both LRRK2 and NOD2 mutations. Finally, we observed a joint effect of LRRK2 and NOD2 variants on cytokine/chemokine secretion with the most significant reduction of secretion observed for the mutant genotypes carried by the twins. These data demonstrated the pleiotropic effects of LRRK2 and NOD2 in response to mycobacterial infection consistent with a role of the identified mutations in the development of early onset leprosy.

**One Sentence Summary:** Variants of *NOD2* and *LRRK2* shared between early onset leprosy, Parkinson’s and Crohn’s disease jointly impact the anti-mycobacterial host response.

## INTRODUCTION

Leprosy is a disease of the skin and peripheral nerves that is caused by infection with *Mycobacterium leprae*. The mode of transmission of leprosy is unknown and humans are the only known medically relevant reservoir of *M. leprae*. Although effective antimicrobial drugs are available, in 2019 over 200,000 new cases of leprosy were detected globally (*1*). The majority of leprosy cases are diagnosed in early adulthood (age 20-40 years) and less than 10% of global cases fall below the 15-year age group (*2*). Even under conditions of high transmission, only 1% of cases are in the 1-4 years’ age group with cases younger than 2 years being exceedingly rare (*2*). This age distribution of cases suggests that prolonged exposure to *M. leprae* or a long incubation period are necessary to result in clinical disease for the majority of exposed persons.

The first genome-wide association study of leprosy identified a striking overlap of genetic risk factors for leprosy and Crohn’s disease (CD), an inflammatory bowel disease (IBD) characterized by a chronic relapsing intestinal inflammation (*3, 4*). When the comparison was extended to the level of risk variants for IBD and excessive inflammatory episodes in leprosy, termed type-1 reactions (T1R), it became apparent that a majority of risk variants were shared between T1R and IBD while a smaller proportion of risk variants were shared between leprosy *per se* and IBD (*5*). A recent study discovered significant pleiotropy of amino acid mutations of the *PRKN* gene for Parkinson’s disease (PD) and T1R while amino acid mutations of the *LRRK2* gene displayed significant antagonistic pleiotropy for T1R and PD (*6*). The same amino acid change protected from leprosy *per se* demonstrating how the same genetic variant can protect from infectious disease while predisposing to host damaging inflammatory responsiveness (*6, 7*). Recently, pleiotropic effects were also observed for amino acid mutations in the *LRRK2* and *NOD2* genes between CD and PD (*8*). In the present study we found that the same *NOD2* and *LRRK2* variants are shared by twins displaying extreme early onset leprosy. We then showed that NOD2 signaling involves LRRK2 and that the LRRK2/NOD2 amino acid substitutions additively impact on immune responsiveness.

## RESULTS

### Whole Genome Sequence analysis

A small nuclear family from Northeast Brazil was identified with leprosy cases in three generations over a two-year period (Fig. 1A). Unusually, monozygotic twins of age <24 months (ID6 and ID7) were both affected by leprosy with nearly identical distribution of skin lesions. The early age of onset for the twins together with the high prevalence of leprosy in the family suggested that host genetic factors may be involved in the familial clustering of the disease. Genomic DNA was obtained for six family members and used for whole genome sequencing (WGS; Fig. 1B). The mean base coverage of all samples was 31±12 fold, with 91.5% of the genomes covered at least 10-fold (Table S1). In total, nearly 8.4 million single nucleotide variants (SNVs) and short indels, including 44,679 coding and splice-site variants, were identified in the six family members. To systematically identify candidate leprosy susceptibility variants, we applied seven variant filtering strategies based on allele frequencies, mode of inheritance and allele distributions within affected family members (Fig. S1). In addition, variant-level and gene-level metrics were used to prioritize variants according to their *in-silico* predicted impact on protein function. In the final screening step, variants were prioritized according to the physiological function of their tagged proteins. Filtering approaches from WGS data that led to the identification of the candidate variants are presented in Fig. S1 and the results are shown in detail in Table S2.

**Fig. 1.**
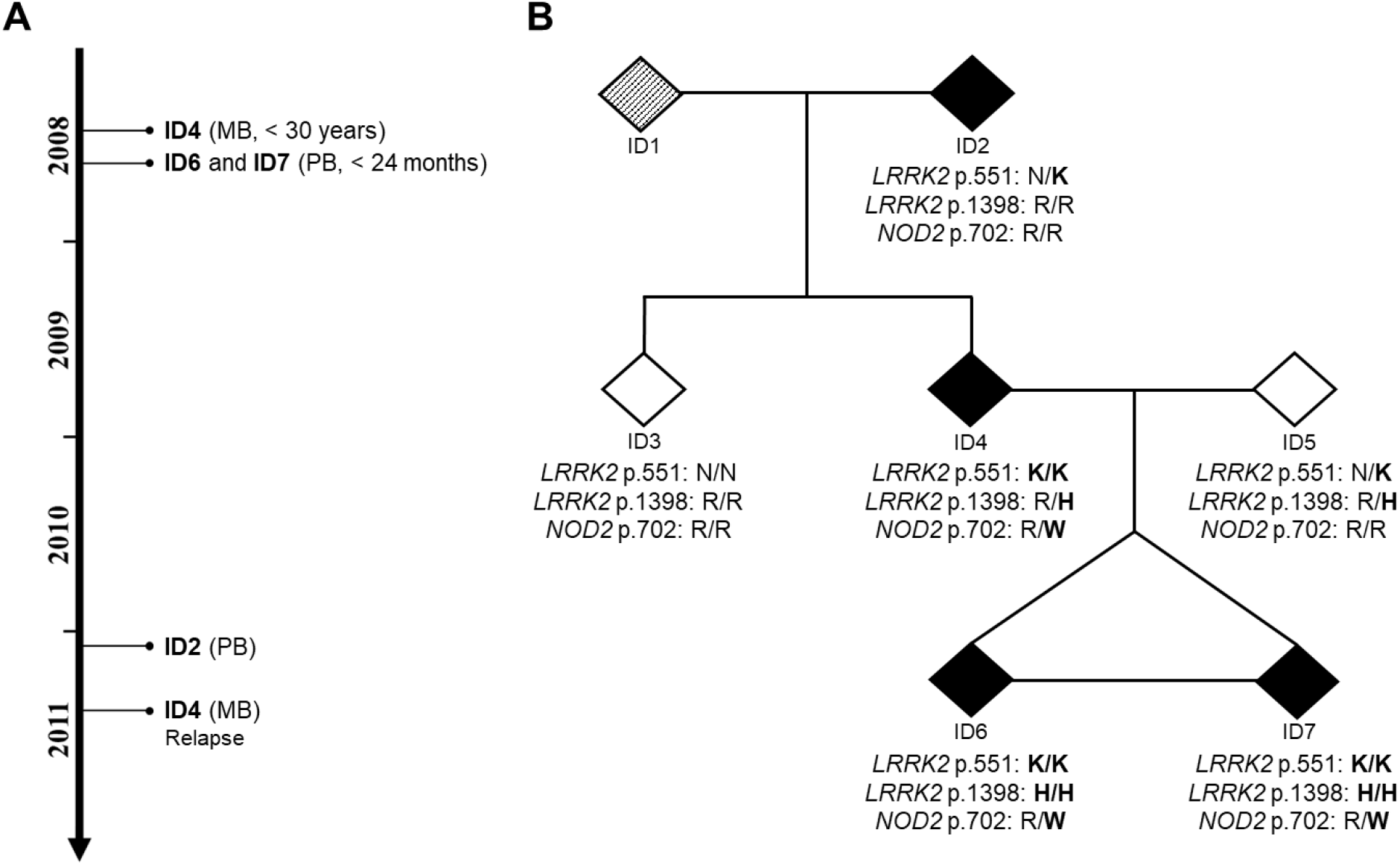
Missense variants in *LRRK2* and *NOD2* detected in a family with early-onset leprosy in monozygotic twins. (**A**) Timeline of leprosy diagnosis among the affected family members, indicating the sample ID, leprosy subtype based on WHO classification and age-at-diagnosis. (**B**) Pedigree of the studied family indicating leprosy phenotype and genotypes of candidate variants *LRRK2* N551K (rs7308720), *LRRK2* R1398H (rs7133914), and *NOD2* R702W (rs2066844) in individuals with whole genome sequencing (WGS) data. For reasons of confidentiality, sex is not indicated in the pedigree. Leprosy patients (regardless of the clinical subtype) are indicated by filled symbols, while an unknown phenotype is indicated by a symbol with diagonal stripes. Monozygosity is represented by a horizontal line linking siblings. PB: Paucibacillary; MB: Multibacillary.

Considering the low prevalence of leprosy and assuming recessive models of inheritance, we selected a minor allele frequency (MAF) of 15% or 30% in any of the HapMap populations as cut-off (Fig. S1). We detected a total of 95 segregating non-synonymous and frameshift indels variants in 51 genes as leprosy risk candidates (Supplementary Methods, Fig. S1 and Table S2). Of these, seven SNPs were predicted *in-silico* to encode protein-damaging variants: rs7271712 (T397M) in *SLC17A9*, rs7308720 (N551K) and rs7133914 (R1398H) in *LRRK2*, rs116409312 (V121L) and rs116063135 (R203H) in *SLC22A24*, rs61740826 (C406Y) in *ZNF678* and rs2229531 (V200M) in *ACP5* (Table S2). The twins and their affected parent (ID4) were homozygous for N551K in *LRRK2*; and only the early-onset twins were compound heterozygous for *SLC22A24* and homozygous for *LRRK2* R1398H as well as *SLC17A9* T397M, *ZNF678* C406Y and *ACP5* V200M (Table S2). Among these variants, *LRRK2* N551K and R1398H had previously been shown to impact on LRRK2 protein activity and are established protective factors for CD and PD (*3, 8*). Conversely, the remaining four genes carrying homozygous missense variants in the twins – *SLC17A9, SLC22A24, ZNF678* and *ACP5* – had no known link to infectious diseases. Hence, the two *LRRK2* variants were considered the top candidates contributing to the early onset leprosy phenotype.

Under dominant models, 37 SNVs and Indels with MAF lower than 10% were identified as leprosy risk candidates, including eight variants prioritized as protein-damaging *in silico* (Fig. S1 and Table S2). Of the genes tagged by novel mutations only the *CR1* gene had prior evidence of a common variant impacting on human infectious disease risk, including leprosy (*9, 10*). The novel *CR1* E1674G amino acid change may deserve further attention in future studies. In addition, one variant (*NOD2* R702W, rs2066844) was a co-dominant risk factor for CD (*11*). A follow-up of *NOD2* variants revealed three additional amino acid changes that had been excluded in the filtering approaches for segregation in the family: P268S, A612T and A725G (Fig. S2). The A725G variant was likely benign and there was no strong evidence for a risk effect of this amino acid mutations in common immune or infectious disease. In contrast, *NOD2* 612T, which was present in the affected parent and grandparent (ID2) but not in the twins, and P268S, which was considered to be too common to have a dominant effect on leprosy risk, had been associated with risk of CD (*8, 12*).

We also searched for deletion structural variants (DSVs) that might contribute to leprosy susceptibility in the family. In the DSV analysis, we identified 55 deletions with length ranging from 0.9 Kb to 43.4 Kb that overlapped coding regions of protein-coding genes. From this, four DSVs passed filtering for segregation in the family (Fig. S1 and Table S3). DSVs overlapping *FSTL4, C18orf32* and *PARVB* were found in three generations of the family where the affected individuals were heterozygous for the variants (Table S3). Only the twins and their affected parent carry the fourth candidate DSV, which overlapped with the *C9orf50* gene (Table S3). There was no known involvement of these genes in either leprosy, infectious or immune-mediated diseases.

Since both *LRRK2* and *NOD2* had been implicated in susceptibility to leprosy, CD and PD, and the corresponding proteins interact *in-vivo* in Paneth cells, we considered these two genes high priority candidates for exerting a joint effect on early onset leprosy susceptibility (*13, 14*). *LRRK2* N551K and R1398H present global MAF of approximately 8.5% (Table S2). However, their frequencies and linkage disequilibrium (LD) pattern vary among populations. In South Asians, both variants are of low frequency (MAF < 5%) and present strong LD (*r*^*2*^ = 1), while in American and African populations they are more common (MAF between 10% and 15%), but with lower LD (*r*^*2*^ = 0.66 and 0.18, respectively). *NOD2* R702W has global MAF of 2.27% (Table S2) and its MAF ranges from 0% in East Asian to 5.1% in European populations. Ancestry estimated based on principal component analysis of the family members of the present study and unrelated individuals from the five HapMap populations suggested a high African (AFR) ancestry for the grandparent ID2 and shared American (AMR) and AFR ancestry for the remaining family members including the twins (Fig. S3). Based on the estimated allele frequencies in the AMR or AFR population and assuming complete LD between the two *LRRK2* variants, the probability of carrying the *LRRK2* and *NOD2* CD and PD-associated alleles by the twins was estimated at approximately 0.08%-0.12% in AMR and 0.007%-0.009% in AFR individuals, which further supported a role of these variants in extreme early onset leprosy.

### LRRK2 variants affect ROS production and apoptosis in response to mycobacteria

To functionally validate the findings from the WGS analysis, we explored the impact of the N551K and R1398H LRRK2 variants on the cellular response to mycobacteria (Fig. 2). The human and mouse LRRK2 proteins are highly conserved with approximately 90% amino acid sequence identity (*15*). Hence, we applied CRISPR/Cas9 technology to knock-in the homozygous candidate LRRK2 variants in mouse RAW264.7 macrophages either individually or as double mutants (DM). A *Lrrk2* knock-out (KO) cell line was also generated as control. The expression of LRRK2 wild-type (WT) and corresponding mutant proteins in RAW264.7 cells is shown in Fig. 2A. Oxidative burst, the generation of reactive oxygen intermediates (ROS), is an important mechanism by which intracellular mycobacterial growth is controlled (*16*). Compared to LRRK2 WT, the LRRK2 R1398H mutation significantly reduced the production of ROS by RAW264.7 cells in response to live *M. leprae* infection (*P* < 0.001 at 1-2h post-infection [p.i.], *P* < 0.01 at 4-6h p.i.) while the N551K variant had no significant effect (Fig. 2B). Interestingly, the LRRK2 DM had a similar effect on ROS production as the R1398H mutation (*P* < 0.001 at 1-2h p.i., *P* < 0.05 at 4h, p.i.), which suggested that the effect of the N551K/R1398H double mutation was mainly due to R1398H (Fig. 2B). Consistent with a previous report, relative to LRRK2 WT expressing cells, the production of ROS was significantly lower in LRRK2 KO cells (*P* < 0.01 at 1h p.i., *P* < 0.001 at 2-6h p.i., Fig. 2B) (*17, 18*). We also investigated if mutant LRRK2 affected ROS production in response to Bacillus Calmette-Guérin (BCG) and we obtained similar results to those in response to *M. leprae* infection (Fig. 2B).

**Fig. 2.**
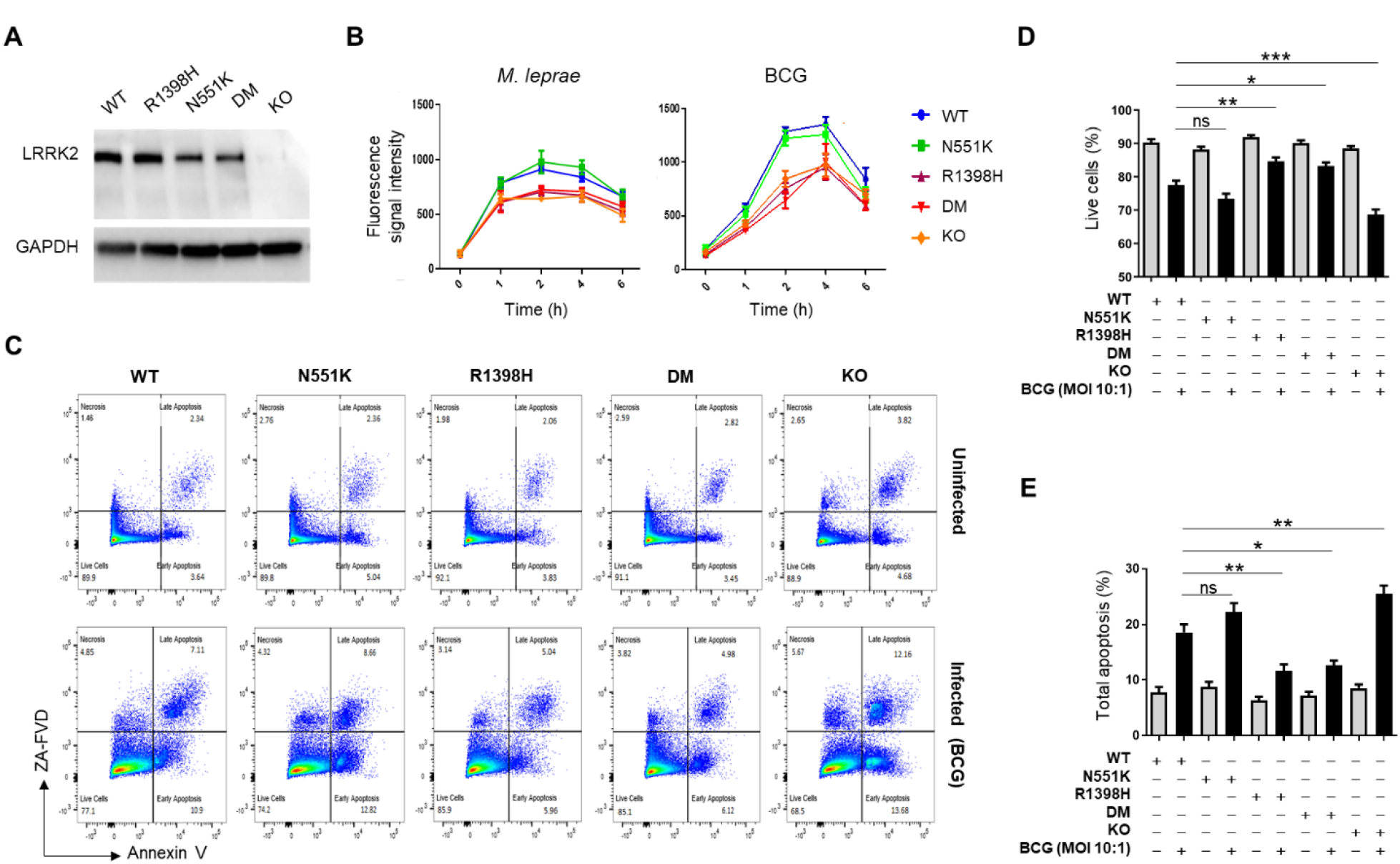
Effects of LRRK2 variants on respiratory burst and apoptosis in response to pathogens. (**A**) Protein levels in RAW264.7 cells of LRRK2 WT, LRRK2 mutants [N551K, R1398H and DM] and no LRRK2 protein (KO). The result is a representative of three independent experiments done by Western blot analysis. The expression of GAPDH is shown as a loading control. (**B**) Kinetics of reactive oxygen species (ROS) production in cells with different *Lrrk2* genotypes after stimulation with live *M. leprae* (left panel) or BCG (right panel). Results present the mean ± SE for three independent experiments (done in triplicates). (**C-E**) Effect of LRRK2 mutations on apoptosis in response to BCG compared to uninfected cells. (**C**) Flow cytometry for apoptosis, which is a representative of two independent experiments (done in triplicates) with similar results. Percentages of live cells (**D**) and total apoptotic cells (**E**) were calculated in uninfected (grey bars) and BCG-infected cells (black bars) expressing different LRRK2 variants. (**D-E**) Data is presented as mean ± SE (n=6). ** 0.001 ≤ *P* < 0.01; * 0.01 ≤ *P* < 0.05.

Apoptosis is part of the innate immune response against mycobacteria (*19*). Mutant LRRK2 variants or loss of LRRK2 expression have been linked to apoptotic cell death (*6, 20, 21*). Hence, we tested if LRRK2 N551K, R1398H or DM affected BCG-induced apoptosis. As shown in Figs. 2C to 2E, in uninfected cells, mutant LRRK2 or absence of LRRK2 protein had no effect on naturally occurring apoptosis by RAW264.7 cells. However, following infection with BCG, apoptosis was significantly increased in LRRK2 KO cells compared to LRRK2 WT cells. In contrast, cells expressing LRRK2 R1398H displayed significantly reduced BCG-induced apoptosis compared to LRRK2 WT cells while LRRK2 N551K had no significant impact on the extent of BCG-induced apoptosis. Finally, expression of LRRK2 DM reduced BCG-induced apoptosis to a similar level as LRRK2 R1398H, suggesting that the effect of the double mutation on BCG-induced apoptosis is mainly due to the R1398H mutation (Fig. 2C to 2E). These results demonstrated how the same LRRK2 amino acid substitution can score as gain or loss of function variant depending on the read-out assay employed.

### Effect of LRRK2 variants and NOD2 R702W on ROS production and apoptosis in response to BCG

Since the twins and their affected parent are heterozygotes for NOD2 R702W (Fig. 1B), we investigated if NOD2 R702W and LRRK2 DM have a synergistic impact on ROS production and apoptosis. Plasmids expressing flag-tagged NOD2 WT, its variant R702W or an empty vector were introduced into RAW264.7 cells carrying LRRK2 WT, LRRK2 DM or not expressing LRRK2 protein (KO). No significant difference in NOD2 WT and NOD2 R702W protein expression was observed across cell lines (Fig. 3A). Transfected cells were infected with BCG (MOI 10:1) and kinetics of ROS production were established. Overexpression of both NOD2 WT and NOD2 R702W proteins increased the ROS production in all three LRRK2 variant cell lines upon infection with BCG (Fig. 3B). When overexpressed in LRRK2 WT cells, NOD2 R702W mediated significantly lower BCG-induced ROS production compared to NOD2 WT (*P* < 0.001 at 2-6h p.i.). When introduced in LRRK2 DM cells mutant NOD2 also mediated a lower induction of ROS (*P* < 0.05 at 2-4h p.i.). However, the R702W effect in LRRK2 WT cells was substantially larger compared to LRRK2 DM cells (Fig. 3B). When overexpressed in LRRK2 KO cells, NOD2 R702W did not significantly reduce ROS production relative to the NOD2 WT (Fig. 3B). Taken together, our results showed that LRRK2 and NOD2 jointly modulated ROS production following BCG infection. Contrary to ROS production, overexpression of NOD2 WT or NOD2 R702W had no significant impact on apoptosis on any LRRK2 background (Fig. 3C). A representative flow cytometry figure of the apoptosis experiments is presented in Fig. S4.

**Fig. 3.**
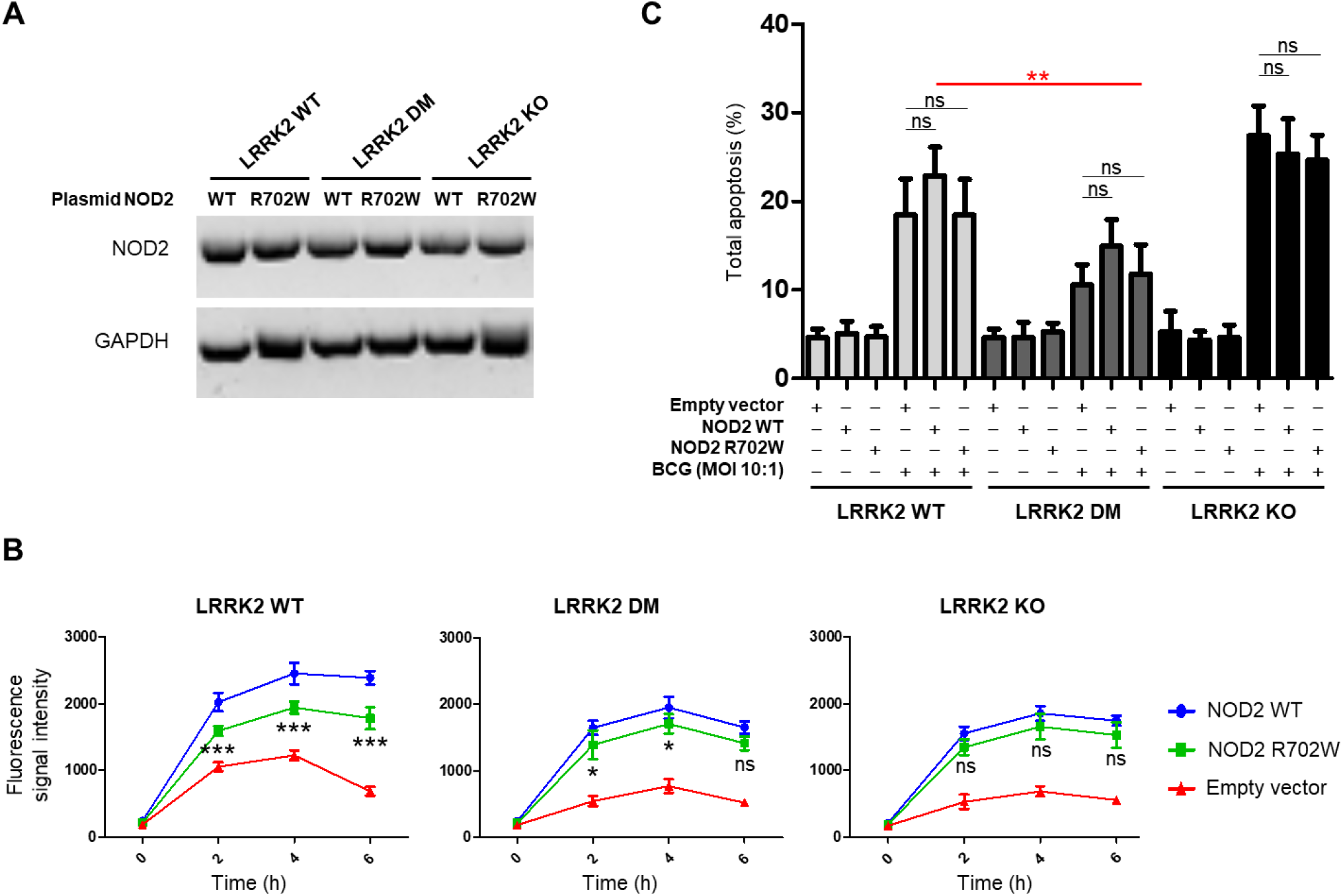
Effects of LRRK2 and NOD2 variants on respiratory burst and apoptosis in response to BCG infection. (**A**) RAW264.7 cells carrying LRRK2 WT, LRRK2 DM and LRRK2 protein (KO) were transfected with plasmids expressing NOD2 WT or mutant NOD2 (R702W). One representative result of three independent experiments is shown. (**B**) Effect of NOD2 WT and R702W on reactive oxygen species (ROS) production after infection with BCG on the background of different LRRK2 genotypes. BCG-induced ROS production in cells transfected with NOD2 WT, NOD2 R702W or an empty plasmid are presented for cell lines with LRRK2 WT (Left panel), LRRK2 DM (middle panel) and LRRK2 KO (right panel). The graphs present one representative experiment (mean ± SD) of three independent experiments, each one done in triplicates. (**C**) Effect of LRRK2 and NOD2 variants on apoptosis in response to BCG. Percentage of total apoptotic cells, including cells with early and late apoptosis, was calculated in uninfected and BCG-infected cells, which is shown in the figure. The illustrated result presents the mean ±SE of two independent experiments (done in triplicates). Significance of difference between LRRK2 WT+NOD2 WT and the genotype carried by the early-onset leprosy twins (LRRK2 DM+NOD2 R702W) is indicated in red. (**B-C**) *** *P* < 0.001; ** 0.001 ≤ *P* < 0.01; * 0.01 ≤ *P* < 0.05; ns: non-significant. BCG: Bacillus Calmette–Guérin; DM: double-mutant; KO: knock-out; WT: Wild-type.

### LRRK2 variants and NOD2 R702W reduces NOD2-dependent RIP2 phosphorylation and NF-κB activity

We used co-immunoprecipitation (Co-IP) to determine if NOD2 interacts with LRRK2 in RAW264.7 macrophages and how the LRRK2 and NOD2 variants segregating in the study family affect this interaction. To test the impact of macrophage activation on the possible LRRK2/NOD2 interaction, we used the NOD2 ligand N-glycolyl muramyl dipeptide (MDP) as trigger (*22*). Co-IP revealed that transfected NOD2 did interact with endogenous LRRK2, and this interaction was independent of the LRRK2 variant and MDP stimulation but sensitive to the NOD2 variant (Fig. 4A). The interaction between LRRK2 and NOD2 proteins was confirmed by co-localization analysis with laser confocal microscopy (Fig. S5). Consistent with Co-IP, these co-localization results demonstrated that NOD2 R702W strongly diminished the interaction between LRRK2 and NOD2 in RAW264.7 macrophages (Fig. S5).

**Fig. 4.**
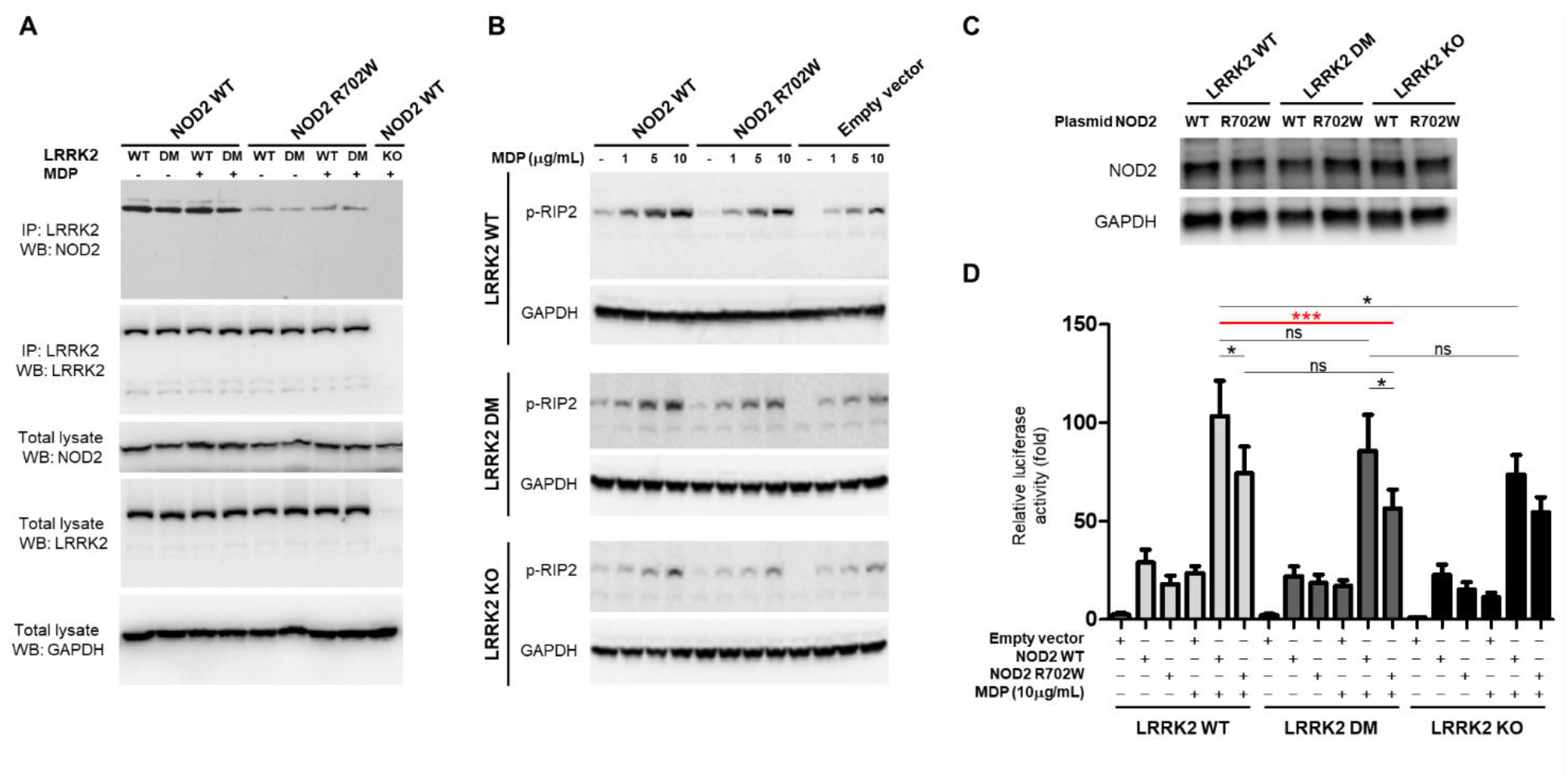
Effects of LRRK2 and NOD2 variants on LRRK2-NOD2 interaction, NOD2-dependent RIP2 phosphorylation and NF-kB activity. **(A)** Protein-protein interaction between endogenous LRRK2 and transfected NOD2 by Co-immunoprecipitation (Co-IP). Twenty-four hours post-transfection, cells were left untreated or treated with N-glycolyl MDP (10 µg/ml) for another 24 hours. Cell lysates were prepared and immunoprecipitated with a rabbit monoclonal antibody against LRRK2. Immunoprecipitants were analyzed by Western blot analysis with antibodies directed against LRRK2 and the FLAG tag of the fused FLAG-NOD2. The expression of LRRK2 and NOD2 using the same antibodies was also analyzed in the total lysate, where GAPDH expression was used as a loading control. LRRK2 KO RAW264.7 cell line transfected with a plasmid expressing NOD2 WT was used as a negative control. (**B**) Effects of NOD2 mutation on MDP-induced phosphorylation of RIP2 at Ser 176 in cells with different LRRK2 genotypes. Twenty-four hours post-transfection, cells were left untreated or treated with different concentrations of MDP for another 24 hours. Phosphorylation of RIP2 (p-RIP2) was analyzed by immunoblotting with a specific antibody against phosphorylated at Ser 176 of RIP2. (**C**) NOD2 expression in the cells from panel B. The displayed results are representative of three independent experiments. The expression of GAPDH is shown as a loading control. (**D**) Effects of LRRK2 and NOD2 variants on NF-κB activity. The three RAW264.7 cell lines with different LRRK2 genotypes were transfected with *NOD2* plasmid (WT, R702W or empty vector) together with NF-κB firefly-Luc plasmid and Renilla-Luc plasmid pRL-TK (internal control). Twenty-four hours after electroporation, cells were left untreated or treated with 10 μg/ml of N-glycolyl MDP for another 24 hours. Cell lysates were subjected to luciferase assays. Results are expressed as relative luciferase activity (fold change), as compared with the luciferase activity of LRRK2 KO cells transfected with empty vector in the absence of N-glycolyl MDP. Significance of difference between LRRK2 WT+NOD2 WT and the genotype carried by the early-onset leprosy twins (LRRK2 DM+NOD2 R702W) is indicated in red. *** *P* < 0.001; ** 0.001 ≤ *P* < 0.01; * 0.01 ≤ *P* < 0.05.

NOD2 activation by MDP results in the activation of the obligate NOD2 kinase RIP2 (alias RIPK2) (*23*). Upon interaction with NOD2, RIP2 becomes auto phosphorylated at two main sites, S176 and Y474. Phosphorylation of RIP2 results in the recruitment of TAK1 and subsequent activation of NF-κB (*23*). We investigated the effect of the NOD2/LRRK2 interaction on RIP2 phosphorylation by Western blot using a RIP2 S176-specific antibody. RIP2 phosphorylation increased after transfection with NOD2 WT and NOD2 R702W constructs in an N-glycolyl MDP concentration dependent manner irrespective of the cellular LRRK2 background (Fig. 4B). However, relative to NOD2 WT, RIP2 phosphorylation mediated by NOD2 R702W was substantially weaker irrespective of the LRRK2 variant carried by the cells even though no significant difference in the expression of NOD2 WT and R702W proteins were observed in these cells (Fig. 4B and 4C). Compared to LRRK2 WT, LRRK2 DM reduced NOD2-driven phosphorylation of RIP2 (Fig. S6). Strikingly, LRRK2 KO resulted in decreased N-glycolyl MDP-induced RIP2 phosphorylation no matter if the cells were overexpressing NOD2 WT or NOD2 R702W (Fig 4B and Fig. S6). Combined, our results identified LRRK2 as important part of the NOD2 signaling cascade and demonstrated that the LRRK2 DM and NOD2 R702W mutations additively reduced RIP2 phosphorylation relative to wild-type proteins.

Next, we performed luciferase reporter assays to detect the combined impact of NOD2/LRRK2 variants on NF-κB activation. Consistent with the effect on RIP2 phosphorylation, N-glycolyl MDP-induced NF-κB activation increased in all three LRRK2 variant expressing cell lines overexpressing NOD2 WT or NOD2 R702W. In both LRRK2 WT and LRRK2 DM cells, NOD2 R702W triggered lower N-glycolyl MDP-induced NF-κB activation compared to NOD2 WT (Fig. 4D). Cells devoid of LRRK2 or expressing LRRK2 DM displayed a strong trend of lower NF-κB activation relative to LRRK2 WT cells for both wild-type and mutant NOD2 (Fig 4D). Consequently, LRRK2 DM and NOD2 R702W had a cumulative effect on the reduction of N-glycolyl MDP-induced NF-κB activation and the variants carried by the early onset leprosy twins reduced NF-κB activation by approximately half compared to the wild-type variants expected in the general population (Fig. 4D).

### Combined effects of LRRK2 DM and NOD2 R702W on cytokine production in response to BCG infection

Next, we asked to what extent LRRK2 DM and NOD2 R702W modulate cytokine/chemokine production. The release of four key mediators (MCP-1, TNF, IL-10, and IL-6) into the supernatant of cell cultures was measured before and after infection with BCG or stimulation with MDP. As expected, infection with BCG induced a stronger cytokine/chemokine response than stimulation with N-glycolyl MDP only (Fig 5). Moreover, the additive effect of NOD2 (WT or R702W variant) overexpression was more pronounced for MDP stimulation. In the absence of transfected NOD2, BCG infection triggered secretion of MCP-1 and TNF while stimulation with N-glycolyl MDP did not, which is consistent with its role as NOD2 ligand. N-glycolyl MDP-triggered secretion of IL-6 remained below the limit of detectability. Release of MCP-1, TNF and IL-6 was substantially reduced by both LRRK2 DM and NOD2 R702W relative to wild-type proteins. There was no significant difference between LRRK2 DM and LRRK2 KO cells, except for the BCG-triggered secretion of MCP-1 which was lower for LRRK2 DM compared to LRRK2 KO (Fig 5A). An unexpected observation was the impact of LRRK2 on the release of the anti-inflammatory cytokine IL-10. RAW264.7 cells expressing LRRK2 DM displayed strongly reduced secretion of IL-10 which was not significantly increased following NOD2 overexpression. Strikingly, absence of LRRK2 resulted in an increased release of IL-10 for both BCG and N-glycolyl MDP stimulation relative to either wild-type or mutant LRRK2-expressing cells even in the absence of overexpressed NOD2 (Fig 5). This observation together with the NOD2-independent impact of LRRK2 mutations on apoptosis activity suggested a pronounced immune regulatory role of LRRK2 (Fig. 2). Collectively, our data show that NOD2 signaling is dependent on the interaction with functional LRRK2. While these observations deserve additional study, the important observation in the context of the present paper was the additive effect of both LRRK2 and NOD2 variants shared by the twins on the release of major mediators of the immune response.

**Fig. 5.**
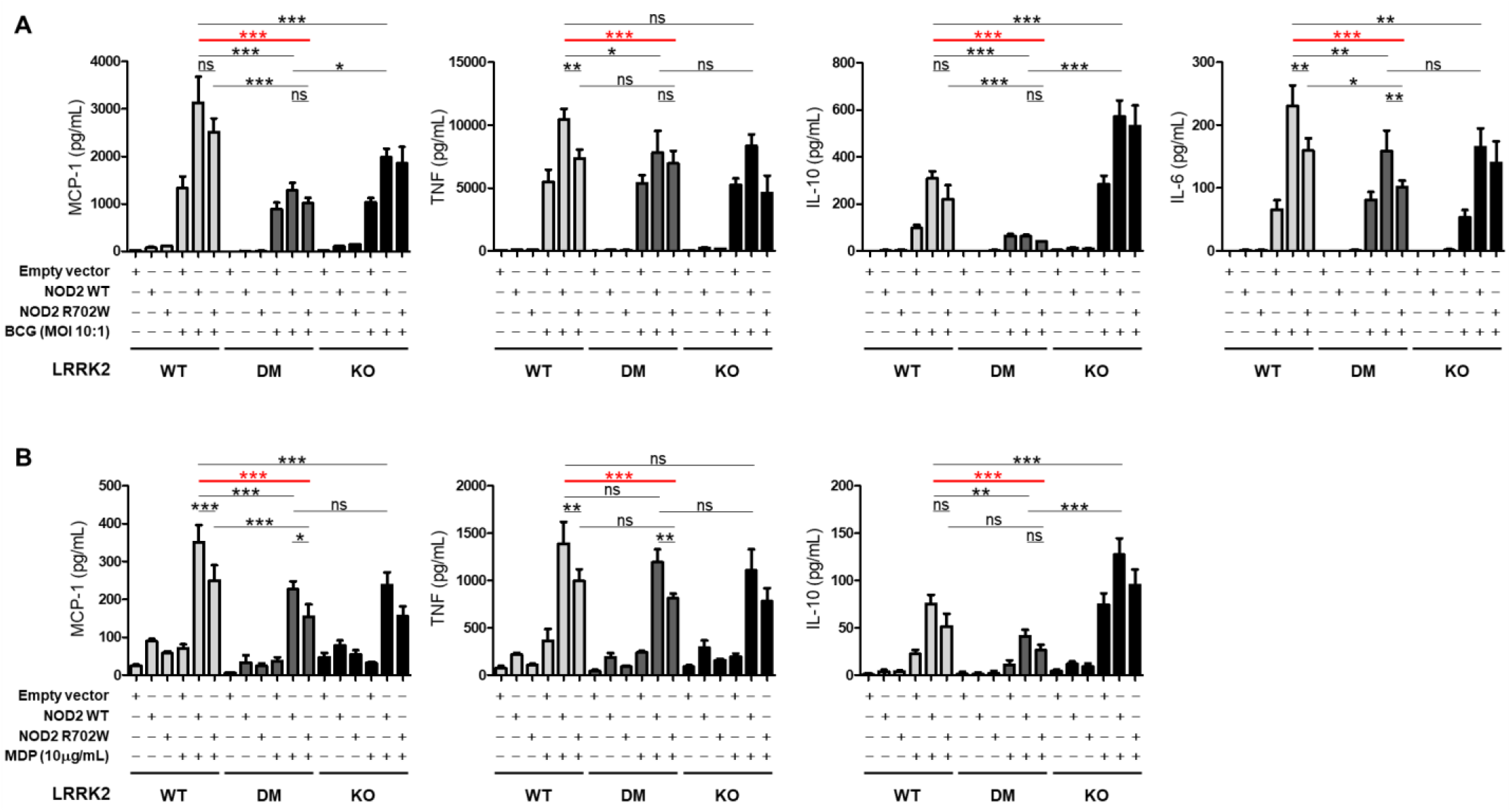
Effects of LRRK2 and NOD2 on cytokine secretion in response to BCG infection or stimulation with N-glycolyl MDP. Cell culture supernatant concentrations for MCP-1, TNF, IL-10 and IL-6 in cells expressing LRRK2 WT, LRRK2 DM (N551K+R1398H) or devoid of LRRK2 (KO) and transfected with NOD2 wild-type (WT), NOD2 R702W or empty vector, on the background of cells. Secreted cytokines were measured following **(A)** infection with live bacillus Calmette–Guérin (BCG) or (**B**) stimulation with N-glycolyl MDP. The concentrations of IL-6 triggered by N-glycolyl MDP stimulation were below the detection limit and are not shown. (**A-B**) Results are presented as mean ± SD of a representative experiment done in triplicate of three independent experiments. Significance of difference between LRRK2 WT+NOD2 WT and the genotype carried by the early-onset leprosy twins (LRRK2 DM+NOD2 R702W) is indicated in red. *** *P* < 0.001; ** 0.001 ≤ *P* < 0.01; * 0.01 ≤ *P* < 0.05; ns: non-significant.

## DISCUSSION

By applying unbiased WGS analysis on a family with monozygotic twins with extreme early-onset leprosy, we identified three coding variants in the *LRRK2* and *NOD2* genes as strong candidates for contributing to early-onset leprosy susceptibility. Rare and common variants in both genes had previously been associated with leprosy and leprosy reactions by candidate and genome-wide association studies (*3, 6, 7, 24-30*). Intriguingly, the same coding variants identified in the leprosy family have been implicated in reduced susceptibility to PD and CD (*8*). Hence, our findings expanded the overlap in the genetic control of these three diseases to specific amino acid substitutions and emphasized intersecting mechanisms of pathogenesis (*5*). By employing a functional follow-up of the genetic findings, our study demonstrated the individual and joint effects of *LRRK2* and *NOD2* on the innate immune response. Of the two *LRRK2* mutations, R1398H, the only *LRRK2* variant homozygous only in the early-onset twins, displayed a predominant functional impact. This was consistent with previous results linking R1398H with increased GTPase activity and Wnt signaling and strengthened the candidacy of R1398H as leprosy susceptibility factor (*8, 31*). While our study was motivated by early onset leprosy, it is likely that the additive interactions of the LRRK2 and NOD2 variants and their effect on key inflammatory host pathways are also valid in PD and CD patients.

*LRRK2* and *NOD2* genes are highly expressed in immune cells in different tissues including the gut, blood and brain (*32*). Here, we decided to probe the impact of the LRRK2 and NOD2 variants on three aspects of anti-mycobacterial host immunity: ROS production, apoptosis, and secretion of immune-modulatory chemokines/cytokines. In addition to being a key effector mechanism in anti-mycobacterial host responses, production of ROS and the resulting oxidative stress are also key events in CD and PD pathogenesis (*33-35*). Apoptosis plays a vital role in host defense against intracellular pathogens, including mycobacteria (*36, 37*). Apoptosis is present in leprosy lesions and may contribute to nerve damage (*38, 39*). Similarly, apoptosis is a key event of IBD pathogenesis and neuronal cell death in PD (*40, 41*). Cytokine responses to *M. leprae* determine the clinical manifestation of leprosy and dysregulation of cytokine-mediated inflammatory host responses has been implicated in PD and CD susceptibility (*42-45*). Importantly, both LRRK2 and NOD2 had previously been implicated in these three investigated host response pathways. LRRK2 modulates apoptotic activity of macrophages following infection with BCG (*6*). Both LRRK2 and NOD2 are modulators of ROS production by immune cells following pathogen challenge (*6, 8, 46, 47*). The loss of pro-inflammatory activity of LRRK2 and NOD2 has been linked to leprosy, PD, and CD, and *LRRK2* enhances NOD2-mediated inflammatory cytokine production (*8, 48, 49*). In the present study, we narrowed down these well-established functional effects of LRRK2 and NOD2 on innate host immunity to specific amino acid variants and their interaction.

While NOD2 signaling and ROS production were significantly modulated by the LRRK2 - NOD2 interaction, apoptosis was not. LRRK2 is an inhibitor of apoptosis and IL-10 secretion, and these were the only readouts where the R1398H mutation presented as gain-of-function. Conversely, in assays that showed an additive effect of NOD2 and LRRK2, i.e. ROS production, N-glycolyl MDP-triggered induction of NF-kB activity as well as N-glycolyl MDP and BCG-triggered pro-inflammatory chemokine and cytokine production, the LRRK2 variants were loss-of-function mutations. While the interaction of LRRK2 and NOD2 was more strongly dependent on the NOD2 R702W mutation compared to the LRRK2 variants, it seemed likely that even in the presence of intact physical interaction, the functional integrity of the complex was reduced. This conclusion was consistent with the impact of the LRRK2 DM on NOD2 signaling, the induction of NF-kB activity and cytokine secretion. Neither the N551K nor the R1398H substitution reduce LRRK2 kinase activity (*8*). Conversely, we had previously shown that the gain-of-kinase activity LRRK2 1618P mutation is a gain-of-function variant for both ROS production and apoptosis (*6*). Taken together these results suggested that LRRK2-mediated ROS production may be dependent of LRRK2 kinase activity while increased apoptosis inhibition is not.

We showed that LRRK2 is an important part of the NOD2 signaling cascade in RAW cells. This finding is consistent with the observation that microglia from *Lrrk2* KO mice displayed a reduced inflammatory response after treatment with α-synuclein pre-formed fibril or LPS (*50*). In a mouse model of colitis induced by dextran sodium sulfate, LRRK2 lies downstream of the β-glucan receptor Dectin-1 and overexpression of LRRK2 leads to the activation of the NF-κB components, TAK1 complex and TRAF6, and the enhancement of pro-inflammatory cytokine secretion (*51*). Co-immunoprecipitation assays revealed an interaction between LRRK2 and NOD2 when both proteins were overexpressed in HEK293T cells. This interaction also occurred between the endogenous LRRK2 and NOD2 in Paneth cells to properly secret antimicrobial peptides into the intestinal lumen and promote gut-microbiota homeostasis (*13*). Moreover, LRRK2 enhanced the phosphorylation of RIP2 at Ser 176 and promoted NF-kB activation, augmenting the production of pro-inflammatory cytokines upon NOD2 activation (*49*). We confirmed and expanded on these previous observations and showed that the LRRK2 and NOD2 mutations synergistically reduced RIP2 phosphorylation and NF-kB activation. Finally, while Ser 176 phosphorylation is essential for RIP2 kinase activity, our results obtained with LRRK2 DM and LRRK2 KO cells confirmed that NF-kB activation may also occur at reduced levels independently of RIP2 phosphorylation (*52, 53*).

As expected from the RIP2 phosphorylation and NF-kB activation experiments, we found that the LRRK2 and NOD2 mutations significantly impaired MCP-1, TNF, IL-6 and IL-10 secretion by RAW cells upon stimulation with BCG or N-glycolyl MDP. The chemokine MCP-1 is involved in recruiting macrophages and monocytes to the sites of infection, and thereby enhances innate inflammatory events. In addition, MCP-1 exerts pleiotropic functions on immune cells, including stimulation of cellular differentiation, proliferation and survival, as well as activating phagocytosis, and efferocytosis (*54*). TNF is a key pro-inflammatory cytokine in mycobacterial infections that triggers granuloma formation while inhibiting mycobacterial growth (*55*). IL-6 plays important role in inflammation and activation of Th1 and Th17 cells. Whereas increased expression of IL-6 is implicated with excessive pro-inflammatory episodes such as T1R, deficiency of IL-6 has been shown to increase susceptibility and bacterial load in animal model of bacterial infection (*29, 56, 57*). IL-10 is an anti-inflammatory cytokine that plays a key role in infections by preventing inflammatory damage to host tissues and a range of pathogens, including mycobacteria, have been shown to manipulate IL-10 signaling to promote survival and infection (*58-61*). Up-regulation of IL-10 expression by IL-27 suppresses IFN-γ-induced antimicrobial activity against *M. leprae* (*62*). IL-10 secreted by different immune cells plays important roles in the progression and phenotype of leprosy (*63-65*). In addition, variant −819 C/T (rs1800871) in the promoter region of *IL10* has repeatedly been found to be associated with susceptibility to leprosy (*66, 67*). All these data suggest an important role of IL-10 in regulation of immune response and even the progression of leprosy. Finally, in our experiments LRRK2 DM as well as LRRK2 KO had a detrimental impact on secretion of pro-inflammatory cytokines by RAW macrophages. However, a recent study of long-term *Mycobacterium tuberculosis*-infected LRRK2 KO mice observed increased transcription of pro-inflammatory cytokines in the lungs of these animals (*68*). These divergent results suggested, as previously pointed out by Shutinoski *et al*, that the outcome of the LRRK2-pathogen interaction may depend on both the pathogen and the length of interaction (*69*).

In previous work, it was shown that the LRRK2 1628P mutation was a risk factor for leprosy but protective for T1R (*6, 7*). The antagonistic pleiotropic effect of the 1628P LRRK2 mutation is a reflection of the two sides of the anti-pathogen host response. Initially a beneficial host response is directed against the infecting pathogen. However, an excessive host response will lead to host cell damage and this host damaging response can occur after the elimination of the pathogen as is the case for leprosy and T1R and other infectious diseases (*70*). Our present data extended the concept of antagonistic pleiotropy of LRRK2 mutations to leprosy and PD/CD. The two mutations were a loss-of-function for BCG and N-glycolyl MDP-triggered cytokine production and also linked to a reduced respiratory burst response and reduced apoptosis. Since leprosy is an infectious disease, a weakened inflammatory host response mediated by the LRRK2 DM and NOD2 R702W is expected to increase susceptibility. Conversely, both PD and CD are inflammatory disorders corresponding to T1R. Given that NOD2 is a microbial sensor and the *LRRK2* and *NOD2* mutation mediated a dampened classical anti-microbial host response this suggested the involvement of microbes in early events of both CD and PD. The latter conclusion is consistent with recent observations in animal models of PD (*69, 71*). Indeed, it is plausible that antagonistic pleiotropy across different diseases does contribute to the maintenance of genetic risk factors in human populations. The results of our study highlight the need for a better understanding of pleiotropy and possible epistatic effect in the dissection of the pathogenesis of common inflammatory disorders.

## MATERIALS AND METHODS

### Study design

In the present study we investigated leprosy genetic risk factors in a three-generational leprosy family, which included monozygotic twins with extreme early-onset disease. WGS was used to identify candidate susceptibility variants considering age-at-diagnosis, models of inheritance, and *in silico* predicted functional impact. Employing this unbiased approach, *LRRK2* N551K and *LRRK2* R1398H (recessive model) and *NOD2* R702W (dominant model) were prioritized as most likely causal variants for extreme early-onset leprosy. Next, follow-up assays were conducted to functionally validate these findings. Genome-editing was used to generate RAW264.7 murine macrophage lines expressing LRRK2 N551K, LRRK2 R1398H, LRRK2 N551K/ R1398H double mutations, or a LRRK2 knock-out (Supplementary methods). ROS production, apoptosis, NF-kB activation and secretion of soluble mediators following *M. leprae*, BCG or N-glycolyl MDP stimulation were compared between these and LRRK2 WT cells. Moreover, to investigate the joint effects of *LRRK2* and *NOD2*, three cell lines expressing *LRRK2* WT, DM or KO were transfected with *NOD2* WT, *NOD2* R702W or empty vector controls. This allowed us to show that *NOD2* and *LRRK2* jointly contribute to ROS production and chemokine/cytokine secretion in response BCG/MDP, and that reduction of this activity was additive for the LRRK2 and NOD2 mutations identified in the twins. As a consequence of this interaction and joint effect, compared to wild-type the genotypes carried by the early onset twins mediated a significantly weaker anti-mycobacterial host response supporting a role of the identified variants in the extreme susceptibility phenotype.

### Studied subjects

A three generational family comprising seven family members – including four leprosy cases – was enrolled from Northeast Brazil. Six family members – or their legal representatives – agreed to participate and signed written informed consent, under protocols approved by local institutional review boards (PUCPR and UFPI). The index case was the affected parent (ID4) who was diagnosed with multibacillary (MB) leprosy in 2008 at the age of <30 years and a second time in 2011. Following the first leprosy diagnosis of the parent, household contact tracing was conducted. As part of this follow-up, both twins (ID6 and ID7) were diagnosed with paucibacillary (PB) leprosy at the age of <24 months by two independent experienced leprologists. In 2011, the grandparent (ID2) was diagnosed with PB leprosy. All individuals in the family had been BCG vaccinated. No patient developed leprosy reactions during treatment and a five-year follow-up. No additional mycobacterial diseases were detected in any family member. Subjects ID3 and ID5 remained unaffected. However, only the unaffected parent ID5 had been in prolonged contact with the three leprosy cases. Therefore, only ID5 was included in the variant filtering approaches as unaffected control.

### Whole genome sequencing

WGS was performed for six family members (ID2 to ID7 from Fig. 1B) on HiSeq® 2500 platform (Illumina) to generate paired-end 150 bp reads at the Genome Quebec and McGill Innovation Centre. Alignment to human reference was done using BWA v0.7.12 (*72*). SNVs and short indels were called using GATK HaplotypeCaller and annotated with wANNOVAR (*73*). Deletion structural variants (DSVs) in autosomal chromosomes were detected using Genome STRiP v2.0 SV pipeline with default parameters (*74*). The sequencing analysis pipeline as well as the strategies for identification of candidate leprosy susceptibility variants are described in detail in the supplementary methods.

### ROS detection

Cells were seeded in 96-well plates at a concentration of 3×10^4^ cells per well and stimulated with IFN-γ (100ng/ml) for 24 hours and then infected with BCG-Russia or *M. leprae* at a MOI of 10:1. At indicated time points following infection, intracellular ROS was detected using ROS-ID total ROS detection kit (Enzo life science) as presented in the Supplementary Methods.

### FACS analysis of apoptosis

LRRK2 WT or CRISPR/Cas9-edited RAW264.7 cells (mock transfected, transfected with *NOD2* constructs or empty vector) were separately cultured at a concentration of 6×10^5^/well in a 6-well plate for 16-18 hrs. The cells were then infected with BCG (MOI 10:1) or left uninfected for 24 hours prior to apoptosis detection. The Annexin V staining was done as detailed by the supplier (Biolegend) and the data was analyzed on FlowJo^®^ v10.4.2 (FlowJo, LLC) with viability and Annexin V single stains as FMOs (See Supplementary Methods).

### Western Blot analysis

Equivalent amounts of total cellular lysates were separated on 4% to 12% Tris-Glycine gels (Invitrogen) and electrophoretically transferred to polyvinylidene difluoride membranes (Millipore, Bedford, MA). The membranes were blocked with 5% BSA in TBS-T (Tris-buffered saline-0.1% Tween 20) for 1h at room temperature (RT), and then followed by incubation with primary antibodies overnight at 4°C. A rabbit monoclonal antibody against LRRK2 (Abcam) was used at 1:1,000 dilution. A mouse anti-GAPDH monoclonal antibody (Thermo Fisher) was used at a 1:10,000 dilution. A mouse monoclonal antibody against FLAG (Sigma) was used at 1: 2,500. A rabbit anti-phospho-RIP2 (S176) (abm) was used at 1:1,000. After incubation, membranes were washed 5 times for 5min with TBS-T and were further incubated with appropriate secondary antibodies coupled to horseradish peroxidase for 1h RT. Upon extensive washing, the membrane was developed with enhanced chemiluminescence detection reagents (Bio-Rad), followed by imaging using a ChemiDoc Touch imaging system (Bio-Rad).

### Co-immunoprecipitation

For co-immunoprecipitations, 24 hours after electroporation, cells were treated with or without 5μg/ml of N-glycolyl MDP for another 24 hours, and then cells were lysed in Pierce IP lysis/wash buffer supplemented with 1X protease inhibitor mixture (Thermo Fisher scientific). Cell lysates were centrifuged at 13,000xg for 10 min to remove cellular debris. Next, 800μg of the total cell lysates was incubated with 5μg of the anti-LRRK2 antibody overnight at 4°C with rotation. Then, 500μl of Pierce protein A/G magnetic beads was added and incubated at room temperature for 1h with rotation. The beads were collected with a magnetic stand and washed three times with IP lysis/wash buffer and once with ultrapure water. The proteins bound to the beads were eluted and analyzed by Western blot. For FLAG immunoprecipitation, cells were lysed in RIPA buffer (50mM Tris-HCI, pH 7.4, 1% NP-40, 0.5% Na-deoxycholate, 0.5% SDS, 150mM NaCl, 2mM EDTA) containing 1X protease inhibitor mixture. Then, 1mg of total cell lysates was incubated with 20μl packed volume of anti-FLAG M2 magnetic beads (Sigma) for 4 hours at 4°C. The beads were washed five times with TBS supplemented with protease inhibitors. The bound proteins were eluted and detected by using Western blot analysis.

### Luciferase Reporter Assays

Luciferase reporter assays were performed using Dual-Luciferase Reporter Assay System (Promega) according to the manufacturer’s instructions. Briefly, 1×10^6^ LRRK2 WT, DM or KO RAW 264.7 cells were suspended in 100μl of a solution (buffer R) containing 10μg of pcDNA3.1+/C-(K)DYK-NOD2, pcDNA3.1+/C-(K)DYK-NOD2 R702W or empty vector control, 3μg of *NF-kB firefly-*Luc plasmid, and 1μg of Renilla-Luc plasmid pRL-TK. Cells were transfected by electroporation using the Neon Transfection System (Thermo Fisher Scientific) at 1,680V for 20 ms and 1 pulse. Cells from each transfection reaction were plated in a 24-well plate with 1×10^5^ cells per well. Twenty-four hours after electroporation, cells were treated with 10μg/ml of N-glycolyl MDP for another 24 hours or left untreated; following this, cell lysates were prepared by using Passive Lysis Buffer (Promega). Luciferase activity was determined from a 20-μL cell lysates and measured on the microplate reader. Firefly luciferase activity was normalized to *Renilla* luciferase activity.

### Cytokine and Chemokine Measurements

For cytokine and chemokine measurements, twenty-four hours after electroporation cells were incubated with or without BCG-Russia (MOI 10:1) for another 24 hours. Cell culture supernatants were collected and centrifuged to remove debris. Cytokines and Chemokines were detected using Milliplex Map (EMD Millipore, St. Charles, MO, USA) multiplex magnetic bead-based antibody detection kits according to the manufacturer’s instructions. A 6-plex premixed kit was used, which included analytes IL-1β, IL-6, IL-10, IL-12 (p40), MCP-1 and TNF. Assay was read using xPONENT 3.1 acquisition software and MAGPIX instrument (Luminex Corporation, Toronto, ON, Canada). Data was analyzed using Mulliplex Analyst software, version 4.2 (EMD Millipore) and presented as picogram of cytokine per milliliter of supernatants (pg/ml).

### Statistical analysis

Statistical analysis was conducted using GraphPad Prism 5 (GraphPad Software, California USA, www.graphpad.com). Kinetics of ROS production were analyzed by comparing the mutant cells with the wild-type cells at each timepoint using two-way ANOVA with Bonferroni correction. In apoptosis, NF-kB activation and cytokine secretion assays, one-way ANOVA and *post hoc* t test with Bonferroni correction was used to compare means between two groups. Adjusted *P* < 0.05 was used as significance threshold, which is represented by asterisks in the figures.

## Data Availability

All data are available in the main text or the supplementary materials.

## LIST OF SUPPLEMENTARY MATERIALS

### Materials and Methods

Fig. S1. Custom filtering approaches for candidate variants identification from WGS data in the studied family.

Fig. S2. All missense variants in *LRRK2* and *NOD2* genes detected by WGS in the studied family.

Fig. S3. Population structure of the studied family using principal component analysis based on 237,150 variants from the 1000G WGS data.

Fig. S4. Flow cytometry of the BCG-induced apoptosis analysis of NOD2-transfected cells expressing different LRRK2 genotypes.

Fig. S5. Representative confocal image of colocalization of ectopically expressed NOD2 with LRRK2.

Fig. S6. Effects of LRRK2 variants on N-glycolyl MDP-induced phosphorylation of RIP2 at Ser 176.

Table S1. Summary of whole genome sequencing data and mapping quality control of six samples from the studied family.

Table S2. Candidate single nucleotide variants (SNVs) and short Indels identified in the studied family by applying the custom filtering approaches shown in Fig. S1.

Table S3. Candidate deletion structural variants (DSVs) identified in the studied family by applying the custom filtering approaches shown in Fig. S1.

Table S4. Oligonucleotides used in the CRISPR/Cas construct of *LRRK2* variants. Reference (*75-88*)

## Acknowledgments

We sincerely thank the family members who agreed to be part of this study. We thank Marcel Behr (McGill University) for the gift of N-N-glycolyl MDP, and J.L. Casanova (Rockefeller University) for comments on earlier versions of the manuscript. We thank Linda Adams and Ramanuj Lahiri from the National Hansen’s Disease Program, HRSA, for the provision of *M. leprae*, which was supported by National Institute of Allergy and Infectious Diseases Interagency Agreement IAA 15006-004. We thank all members of the Schurr lab at the MUHC-RI and the Mira lab at PUCPR for useful comments and suggestions to the study and writing of the manuscript. We also thank members of França da Costa lab at UFPI, the Probst lab at ICC as well as Kai Sheng for technical support. We thank Min Fu and Shi-Bo Feng for technical support and the RI-MUHC Molecular Imaging Platform for infrastructure support on the use of the confocal microscope.

## Funding

Canadian Institutes of Health Research (CIHR) grant FDN-143332 (ES).

Conselho Nacional de Desenvolvimento Científico e Tecnológico (CNPq) grant Universal/2011 (MTM).

Conselho Nacional de Desenvolvimento Científico e Tecnológico (CNPq) grant Productivity (PQ) 304368/2018-0 (MTM).

MD-S was supported by a fellowship (3407/15-2) from Coordenação de Aperfeiçoamento de Pessoal de Nível Superior (CAPES), Ministry of Education of Brazil.

JM and AA are supported by Laboratoire d’Excellence Integrative Biology of Emerging Infectious Diseases grant ANR-10-LABX-62-IBEID, the Investments for the Future program grant ANR-10-IAHU-01 and MYCOPARADOX ANR project grant ANR16-CE12-0023.

National Institute of Allergy and Infectious Diseases Interagency Agreement IAA 15006-004 for provision of live *M. leprae*.

Cedar (WestGrid) and Guillimin (Calcul Québec) high performance computing clusters by Compute Canada (ES).

## Author contributions

Conceptualization: ES, MTM, YZX, MD-S,

Formal Analysis: YZX, MD-S

Investigation: YZX, MD-S, ALFC, ST

Visualization: YZX, MD-S

Funding acquisition: ES, MTM

Supervision: ES, MTM

Resources: ALFC, WC-M, JM, LA, AA, AC, VMF, CMP, MTM, ES

Writing – original draft: YZX, MD-S, ES

Writing – review & editing: YZX, MD-S, ES, MTM, VMF, AC, JM, LA

## Data and materials availability

All data are available in the main text or the supplementary materials.

