## Supplementary materials for "Early onset leprosy reveals a joint effect of *LRRK2* and *NOD2* variants"

#### Supplementary methods

##### Sample collection from the family

Genomic DNA was extracted from whole blood by salting out method (*75*). The extracted genomic DNA was quantified using Qubit® dsDNA BR Assay in Qubit® 2.0 Fluorometer (Thermo Fisher Scientific). DNA samples with a 260/280 nm ration ranging from 1.5 to 2.0 were considered adequate.

##### Whole genome sequencing data analysis

Quality assessment of the raw data was performed using FastQC v0.11.4 software (Babraham Bioinformatics; http://www.bioinformatics.babraham.ac.uk/projects/fastqc/). The reads were mapped to human genome reference GRCh37+decoy using the BWA-mem algorithm on BWA v0.7.12 (*72*). Mapped reads were sorted according to their genomic coordinate position and PCR duplicates were flagged using Picard v1.134 (Broad Institute, GitHub Repository; https://github.com/broadinstitute/picard). Next, local realignment around indels and base recalibrations were performed using GATK v3.5 (*76*). Quality assessment of the mapped reads was performed using QualiMap v2.1.1 (*77*). GATK HaplotypeCaller was used to call SNVs and short indels for each sample, followed by GenotypeGVCFs for the six samples together. Variant Quality Score Recalibration from GATK, using default parameters was used to reduce the amount of false positive (*78*). Genotypes with GQ lower than 20 were removed. Finally, the variants were annotated using wANNOVAR (2016) (*73*).

Deletion structural variants (DSVs) in autosomal chromosomes were detected using Genome STRiP v2.0 SV pipeline with default parameters (*74*). For that, a total of 25 high-coverage WGS samples from the 1000G database were included in this step to run together with the 6 samples from the studied family (*79*). Next, GATK was used to apply hard filtering to remove low quality deletions as follow: GSELENGTH $< 200$; GSCLUSTERSEP $\leq 2.0$ or NA; GSM1 $\leq 0.5$ or $\geq 2.0$ or NA; GLINBREEDINGCOEFF $< -0.2$; GSNONVARSCORE $\geq 13.0$; GSDUPLICATESCORE $\geq 0$ or DOSAGE_CORRELATION $\geq 0.5$; call rate $\geq 80\%$. Overlap of the DSVs to protein-coding genes was annotated using GeneOverlap command on Genome STRiP.

To identify the population structure of the studied family, principal component analysis (PCA) was performed using PLINK v1.9 (*80*). For that, genotypes from VCF files were converted to PLINK format using BCFtools (http://github.com/samtools/bcftools). Autosomal variants were pruned based on LD (window size of 50kb, step size of 5 and variance inflation factor of 1.5) and MAF > 10% as implemented in PLINK. Variant pruning was done using the samples from the 1000 Genomes Consortium (1000G) representing the five super populations worldwide: African/African American, Admixed American/Latin, East Asian, South Asian and European (*81*). From the 428,824 pruned variants, A/T and C/G SNPs and variants absent from the studied family were excluded. In total, 237,150 variants were used for PCA analysis including 2,504 unrelated individuals from 1000G and the six family members from the present study.

**Candidate variant detection from WGS data**

Seven custom filtering approaches were applied to help identify candidate variants (Fig. S1). The filtering steps were based on i) the variant location within protein-coding genes (coding or splice-site variants), ii) type (missense, nonsense, frameshift indels or splice-site variant), iii) variant frequency in population samples from the database, iv) inheritance model (dominant or recessive) and v) age-at-diagnosis of the affected family. To apply filtering step 3, we searched the variant frequency in the following population samples from the 1000G and Exome Aggregation consortium (ExAC) databases: African/African American, Admixed American/Latin, East Asian, South Asian and European (Non-Finnish European from ExAC) (*81, 82*). Different thresholds of variant frequencies were used in the four approaches following the recessive model (Filtering approaches #1 to #4 in Fig. S1), while in the dominant model, we selected variants that were not reported in the 1000G nor ExAC databases (Filtering approaches #5 to #7 in Fig. S1). Based on the age-at-diagnosis of the leprosy patients in the family (Fig. 1A), in filtering step 5 we searched for variants present in i) all the affected family members, regardless of the age-at-diagnosis (Filtering approaches #1 and #5 in Fig. S1); ii) the affected parent and the twins, which are the cases younger than 30 years (Filtering approaches #2 and #6 in Fig. S1) and iii) only in the twins, that are the early-onset cases in the family (Filtering approaches #3, #4 and #7 in Fig. S1). In addition, variants in genes previously associated with leprosy were analyzed in more detailed. For that, Huge Navigator database was used for searching entries related to leprosy phenotypes up to February 2016 (*83*). Filtering steps were applied to identify candidate variants in these genes as implemented in the WGS data, but with a less stringent threshold for variant frequency in databases (Fig. S1). For known leprosy genes, thresholds of MAF < 50% and MAF < 10% in the four populations from 1000G/ExAc were used in the recessive and dominant models, respectively (Fig. S1).

Once candidate variants were identified, variant-level and gene-level metrics based on computational prediction were used to prioritize the variants that are most likely to have an impact on the protein structure and function. For that, we used PolyPhen-2 v2.2.2r398 and CADD v 1.4 as variant-level metrics, and GDI (2016) as gene-level metric (*84-86*). We focused on variants that presented the three following criteria: i) it has scaled CADD score ≥ 20 (Top 1% most deleterious), ii) it is a missense variant with PolyPhen-2 HumVar score > 0.446 (possibly or probably damaging), a nonsense variant, frameshift indel or splice-site variant and iii) it is located in a gene with GDI score < 13.84 (medium or low GDI). Among compound heterozygous variants (Filtering approach #4 in Fig. S1), we prioritized genes with GDI score < 13.84 where both variants reached criteria one and two. Linkage disequilibrium (LD) estimates between *LRRK2* N551K and R1398H were performed using Haploview software v4.2 based on genotyping data from the five populations from 1000G (*81, 87*).

For detection of candidate DSVs, deletions reported as overlapping with exon, CDS or gene were selected. Then, we applied the same filtering approaches as used for SNV and short indels for the recessive and dominant model (Fig. S1). DSV with call-rate < 80% in the 31 samples were excluded. We searched whether the candidate deletions were present on Database of Genomic Variants (DGV) catalog and kept those that were found in less than 50% of the sequenced samples from 1000 Genomes database (Fig. S1) (*81, 88*).

**Whole exome sequencing**

Whole exome sequencing (WES) in five family members (ID2 and D4 to ID7 from Fig. 1B) was used as a validation of candidate SNVs and short Indels detected from WGS analysis. Exome was captured using the Targetseq Exome kit (Thermo Fisher Scientific). To cover the target regions, this in-solution array contains more than 2 million oligonucleotide probes ranging from 60 to 100 bp that tile 52.7 Mb of target regions including the exome and flanking areas. Then, the exon-enriched DNA libraries were sequenced by 200 bp single-end reads on Ion Proton™ Sequencer (Thermo Fisher Scientific) using the Ion PI™ Chip v2 (Thermo Fisher Scientific). Quality assessment of the raw data was performed using FastQC software. Sequence data analysis was conducted using a pipeline for variant discovery with Torrent Suite (TS) software v5.0 available on GitHub (https://github.com/iontorrent/TS). Using TS, reads were aligned to the human reference using map4 command line implemented on TMAP (https://github.com/iontorrent/TS/tree/master/Analysis/TMAP). Mapped reads were sorted according to their genomic coordinate position using SortOrder command and PCR duplicates were flagged with MarkDuplicates in Picard. Quality assessment of the mapped reads was performed using QualiMap. Variant calling was performed with Torrent variant caller (TVC) plugin from TS software, using “Germline - Proton TargetSeq - High stringency” parameter option with default settings (https://github.com/iontorrent/TS/tree/master/plugin/variantCaller). Identification of single nucleotide variants (SNV) and dinucleotide variants (DNV) was performed in regions with coverage $\geq$ 10X, while indel calling was performed only in regions with coverage $\geq$ 20X. The lists of variants from all the samples were combined in one multi-samples VCF file using CombineVariants tool in GATK. Finally, genotypes of the candidate variants from the WGS filtering approaches were compared between WGS and WES for variants detected by both methods (Table S2).

**Genome-editing with CRISPR/Cas9**

1. Synthesis of gRNAs: The gRNAs for generation of LRRK2 N551K, R1398H were synthesized by using GeneArt precision gRNA synthesis kit (Thermo Fisher) according to the manufacture's instruction. Prior to making gRNAs, 34-nucleotide forward and reverse target DNA oligonucleotides were designed using the CRISPR search and design tool (Thermo fisher) and synthesized (Table S4). Then the gRNA DNA templates were PCR assembled and gRNAs were synthesized by *in vitro* transcription. The gRNAs were purified and their concentrations were measured. TrueGuide™ Synthetic sgRNA for generation of *Lrrk2* KO cell line was purchased from Thermo Fisher (Assay ID: CRISPR206078_SGM).

2. Electroporation: One day prior to transfection, RAW264.7 cells were split into a new flask with fresh growth medium such that the cells reach 70-90% confluent the following day. On the day of electroporation, cells were washed with PBS (without $Ca^{2+}$ and $Mg^{2+}$), digested with 0.25% trypsin-EDTA for 8-10 min at 37°C. After neutralization with growth medium, the cells were counted and appropriate amounts of cells ($1 \times 10^5$ cells per transfection) were transferred to a 1.5 ml microcentrifuge tube. The cells were washed once with PBS by centrifugation at 500g for 5 min. At the same time as preparation of cells for electroporation, 2µg Cas9 protein and 400 ng gRNA were mixed in 10 µl of resuspension buffer R and incubated at room temperature for 10 min. Prepared cells were re-suspended in the buffer R containing Cas9-gRNA complex and 50 pmol of donor HDR templates (Table S4) was added. Cell mixture was transferred into a 10 µl Neon tip with Neon pipette and electroporation were performed using the parameters as following:

pulse voltage 1680 V, pulse width 20 ms and pulse number 1. After electroporation, cells from two Neon tips were immediately mixed into prewarmed 1 ml growth medium in a well of 12-well plate and cultured for 4 days.

3. Restriction fragment length polymorphism (RFLP) assay: Genomic DNA was extracted from RAW264.7 cells transfected with Cas9-gRNA and donor HDR templates. Genomic DNA was then PCR amplified with primers flanking the donor target region (see Table for PCR primer sequences). The amplification was carried out with AmpliTaq Gold 360 master mix (Thermo Fisher), using the following cycling condition: 95 °C for 10 min for initial denaturation; 40 cycles of 95°C for 30s, 60°C for 30s and 72°C for 35s; and a final extension at 72°C for 7 min. Then, 1 μg PCR products were digested with 10 U of BstUI at 60°C or AvaI at 37°C overnight and resolved on 1.2% agarose gel.

4. Single cell clone analysis: Single cells were prepared by digestion of cells with 0.25% trypsin-EDTA. Cells were counted and serially diluted to 2 x $10^4$cells/ml, 5 x $10^2$cells/ml and 5 cells/ml. Next, 200 µl of 5cells/ml was dispensed to each well of 96-well plates using a multichannel pipette. Plates were incubated at 37°C in a 5% $CO_2$ incubator.

5. Screening knock-in mutation (N551K and R1398H): Genomic DNA was isolated from single clones. The donor target region was PCR amplified with AmpliTaq Gold 360 master mix (Thermo Fisher) (see Table S4 for PCR primer sequences). PCR amplicons were sequenced using standard Sanger sequencing.

6. Screening KO: Cell lysates were prepared from single cell clones and western blot analysis were used to screen knockout clones.

#### Cell Culture and NOD2 Transfection

LRRK2 WT, LRRK2 DM or LRRK2 KO RAW264.7 cells were maintained in DMEM medium supplemented with 10% fetal bovine serum and 1% streptomycin-penicillin, and incubated in a humidified atmosphere containing 5% $CO_2$ at 37°C. The cells were passaged every 3 days. Plasmid pcDNA3.1+/C-(K)DYK-NOD2 encoding Flag tagged human wild-type NOD2 was purchased from GenScript. A plasmid and pcDNA3.1+/C-(K)DYK-NOD2 R702W encoding NOD2 R702W variant was generated using a QuickChange XL site-directed mutagenesis kit (Agilent Technologies) according to the manufacturer's instructions. Primers used for generation of NOD2 R702W mutation were 5'-ggcctggcgccagagcagggcct-3' and 5'-aggccctgctctggcgccaggcc-3'. The NOD2 R702W mutation was confirmed by Sanger sequencing. Plasmids pcDNA3.1+/C-(K)DYK-NOD2 , pcDNA3.1+/C-(K)DYK-NOD2 R702W and pcDNA3.1+/C-(K)DYK empty vector (negative control) were transfected into the three RAW264.7 cells lines using a Neon transfection system (Thermo Fisher Scientific). Briefly, 1 × $10^6$ cells were suspended in 100 μL of buffer R containing 15 μg of plasmid and electroporated at 1,680 V for 20 ms and 1 pulse.

#### Preparation of BCG-Russia and *M. leprae*

BCG Russia culture was maintained in middlebrook7H9 medium supplemented with 10% ADC, 0.1% Tween 80, and 0.2% glycerol at 37°C on a roller. On the day of infection, appropriate amount of log-phase BCG-Russia were transferred to a 50 ml conical tube and pelleted by centrifugation at 2500 rpm for 6 min. The supernatant was removed, and the cell pellet was washed once with 1x PBS and re-suspended in complete DMEM medium. To break up large aggregates into single cells, the re-suspended BCG was treated in water bath sonication for 20s x 5 times, followed by passing the BCG through a 22 1/2-G needle 8 times. The remaining bacterial clumps were removed by centrifugation for 5min at a centrifugation force of 100g. Bacterial load was determined by plating serial 10-fold dilutions of BCG on Middlebrook 7H10 agar plate (supplemented with 10% OADC) and counting colonies after incubation for at least 3 weeks.

Viable *M. leprae* was obtained from the National Hansen's Disease Program, Health Resources and Services Administration, Baton Rouge, LA, USA.

**ROS detection**

Intracellular ROS was detected using ROS-ID total ROS detection kit (Enzo life science) according to the manufacture's instruction. For that, cells were carefully washed with 200 µl/well of 1× wash buffer. Following wash buffer removal, 100 µl/well of ROS detection mix (4 µl of 5mM oxidative stress detection reagent /10 ml of 1× wash buffer) was added prior to incubation of plates in a humidified incubator (37°C, 5% $CO_2$) for 30 min and reading were acquired at wavelength 488/520nm on a plate reader. The experiment was performed three times, each in triplicate.

**FACS analysis of apoptosis**

The Annexin V staining was done according to the manufacture's instruction (Biolegend). Briefly, the cells were first detached from the culture plates and washed twice with 2mL of 1X azide-free and serum/protein-free PBS at RT. The supernatant was discarded. Then, 0.5µl of Zombie Aqua™ fixable viability dye (ZA-FVD) was added to 100µL of cells in 1X azide-free and serum/protein-free PBS and incubated in dark for 30 minutes at 2°C. After incubation, the cells were washed twice with 1X azide-free PBS+0.2% BSA. The cells were washed once with

1X azide-free PBS+0.2% BSA and then once with 1X Annexin V Binding Buffer (BD Biosciences). The cells were resuspended in 1X Binding Buffer at $2x10^6$ cells/ml. Next, 5μl of Annexin V-APC were added to 100μL of the cell suspension and incubated in dark for 15 minutes at room temperature. After incubation, the cells were washed twice with 2ml of 1X Binding Buffer. The supernatant was discarded. The cells were resuspended in 200μl of 1 X Binding Buffer and immediately collected by flow cytometry with BD FACSCanto II (BD Biosciences). The data was analyzed on FlowJo® v10.4.2 (FlowJo, LLC) with viability and Annexin V single stains as FMOs.

**Co-localization with laser confocal microscopy**

Transfected or untransfected cells were seeded in 8-well chambers. Twenty-four hours after transfection, cells were treated with or without 5 μg of N-glycolyl MDP 2 for another 24 hours. Cells were fixed with 4% paraformaldehyde, permeabilized with 0.1% Triton X-100. Samples were blocked with blocking buffer [5% BSA, 2.52 mg/ml glycine in PBST (PBS + 0.1% Tween 20)] and incubated with the primary antibodies diluted in blocking buffer – rabbit anti-LRRK2 1:500 and mouse anti-FLAG 1:250 (for NOD2) – overnight at 4°C. Cells were washed with PBS for 3 x 5 minutes and incubated with secondary antibodies prepared in a diluted (1:5) blocking buffer [Alexa Fluor 488 goat anti-mouse IgG (H+L) 1:1000 and Alexa Fluor 594 donkey anti-rabbit IgG (H+L) 1:1000] for 1 hour. Cells were washed with PBS for 3 x 5 minutes and nuclei were stained with DAPI. Images were obtained by confocal microscopy. Colocalization between LRRK2 and NOD2 was measured from 25-30 cells by Zeiss 2012 ZEN confocal software.

### Supplementary Figures

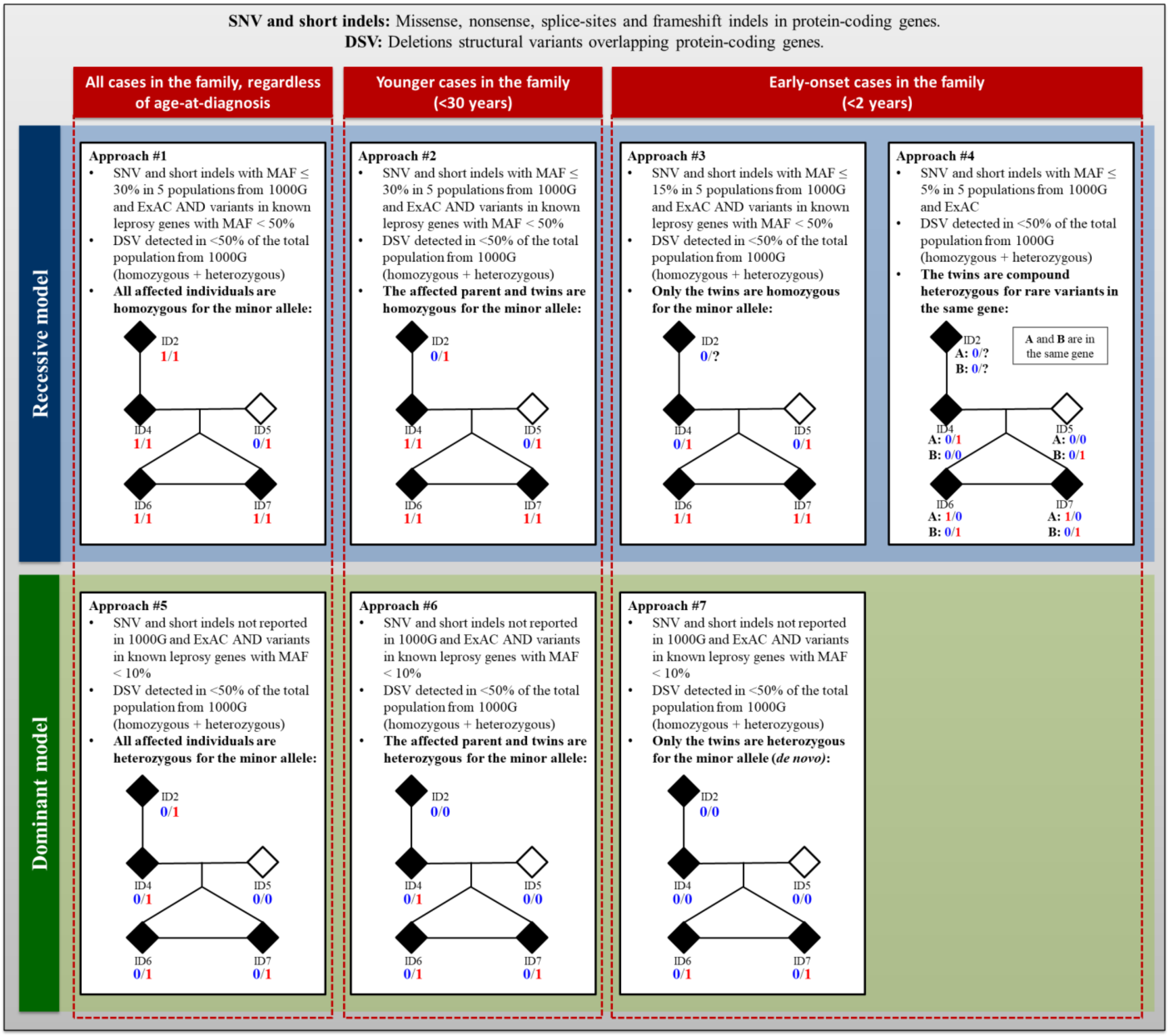


**Fig. S1. Custom filtering approaches for candidate variants identification from WGS data in the studied family.** First, variants were selected based on their location and impact in protein-coding genes (shown on top). Then, seven different filtering approaches were applied (approaches #1 to #7). These filtering steps were based i) on the variant frequencies in public databases, ii) on the model of inheritance and iii) on the age-at-diagnosis of the leprosy-affected family members. Specifically, recessive (#1 to #4)

and dominant (#5 to #7) models were tested based on the presence of the variant in all affected family members (#1 and #5), only in the cases younger than 30 years (#2 and #6) and only in the early-onset twins with less than 2 years (#3, #4 and #7). In the pedigree, men and women are represented by boxes and circles, respectively. Leprosy patients, regardless of the subtype, are indicated by filled symbols. Monozygocity is represented by a triangle. The number zero in blue represents the reference allele and the number one in red corresponds to the variant. The sample ID is the same as Fig. 1. The lists of candidate variants detected using these approaches are presented in Table S1 and Table S2. 1000G: The 1000 genome consortium database; DSV: deletion structural variant; ExAC: The Exome Aggregation consortium database; Indel: insertion/deletion; SNV: single nucleotide variant.

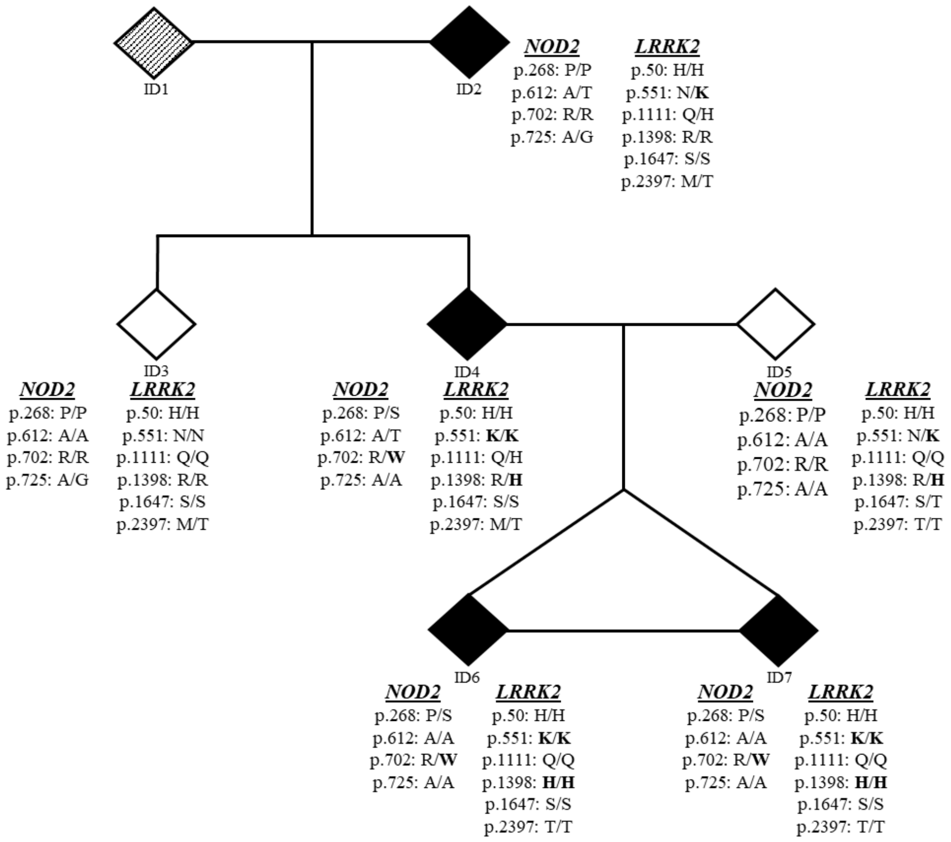


**Fig. S2. All missense variants in *LRRK2* and *NOD2* genes detected by WGS in the studied family.** The *LRRK2* missense variants found in the family were rs2256408 (R50H), rs7308720 (N551K), rs78365431 (Q1111H), rs7133914 (R1398H), rs11564148 (S1647T) and rs3761863 (M2397T). Four missense variants were detected in *NOD2*, which were rs2066842 (P268S), rs104895438 (A612T), rs2066844 (R702W) and rs5743278 (A725G). Among these variants, *LRKK2* N551K and R1398H passed filtering approaches #2 and #3, respectively; while *NOD2* R702W passed filtering approach #6 (see filtering approaches in Fig. S1). The alternative allele from the three candidate variants that passed filtering are shown in bold. No coding indels were detected in *LRRK2* and *NOD2* genes in the WGS data from the studied family. For reasons of confidentiality, sex is not indicated in the pedigree. Leprosy patients, regardless of the subtype, are indicated by filled symbols, while unknown phenotype is indicated by symbol with diagonal stripes. Monozygosity is represented by a horizontal line linking siblings. The sample ID is the same as Fig. 1.

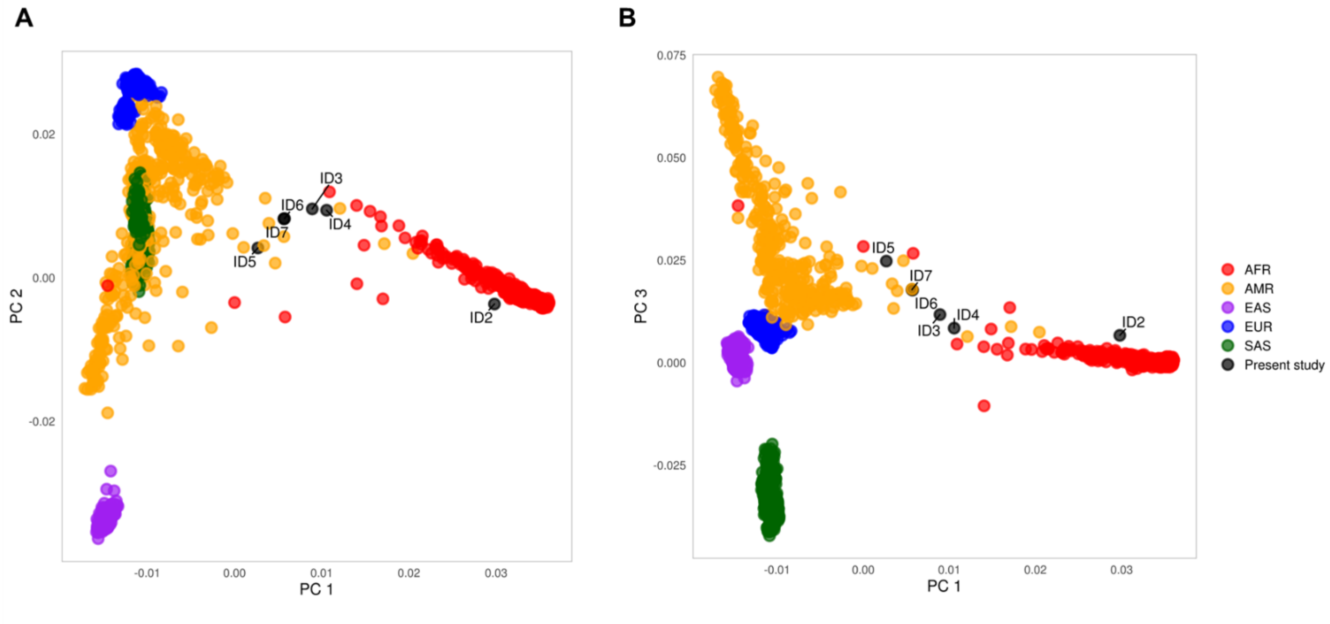


**Fig. S3. Population structure of the studied family using principal component analysis based on 237,150 variants from the WGS data.** Each dot represents an individual, including the six family members from the present study and 2504 unrelated individuals from the 1000 Genomes Consortium representing the five super populations: African/African American (AFR), Admixed American/Latin (AMR), East Asian (EAS), European (EUR) and South Asian (SAS). The sample ID is the same as Fig. 1. (**A**) First and second components are plotted on the *x* and *y* axis, respectively. (**B**) First and third components are plotted on the *x* and *y* axis, respectively.

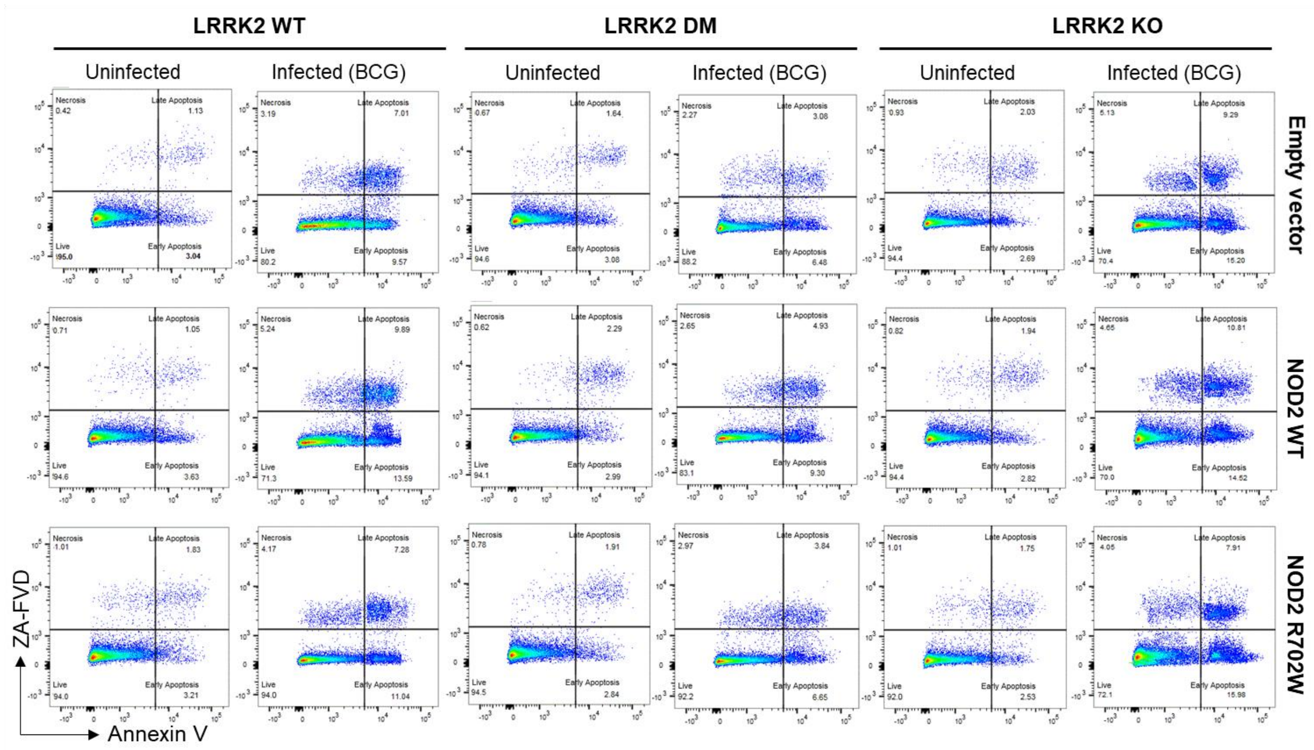


**Fig. S4. Flow cytometry of the BCG-induced apoptosis analysis of NOD2-transfected cells with different LRRK2 genotypes.** LRRK2 Wild-type (WT), CRISPR/Cas-edited LRRK2 double-mutant (DM, N551K+R1398H) and LRRK2 knock-out (KO) RAW264.7 cell lines were transfected with plasmids expressing NOD2 WT, NOD2 mutant (R702W) or an empty vector [pcDNA3.1+/C-(K)DYK] as a control. Twenty-four hours post-transfection, cells were left uninfected or infected with live bacillus Calmette–Guérin (BCG)-Russia (MOI 10:1) for another 24 hours. Cells were then harvested, stained with Annexin V/ZA-FVD, and analyzed by flow cytometry for apoptosis. The illustrated result is a representative of two independent experiments with similar results (done in triplicates).

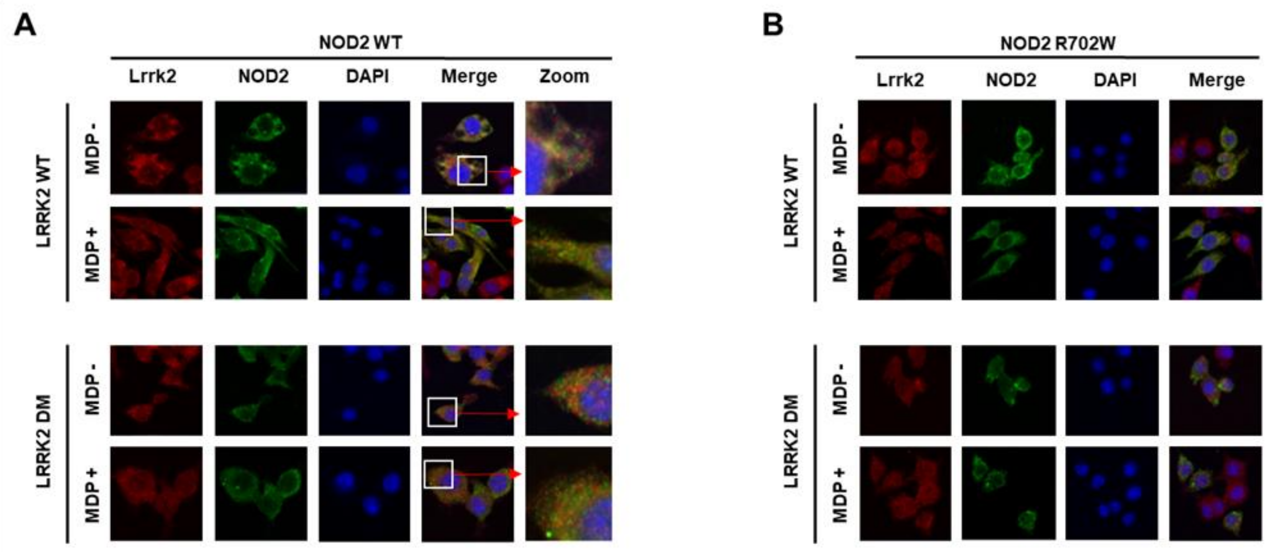



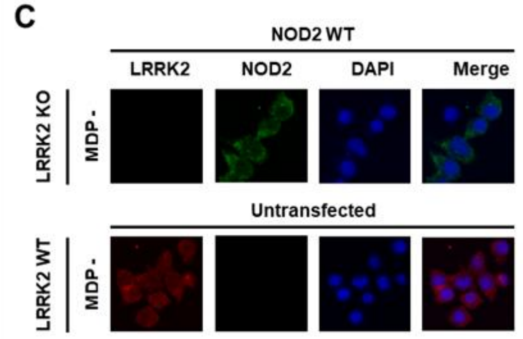



**D**

Co-localization of LRRK2 with NOD2 [mean % (± SD)]

| | NOD2 WT | | NOD2 R702W | |
|---|---|---|---|---|
| | MDP - | MDP + | MDP - | MDP + |
| **LRRK2 WT** | 33.36 (± 3.90) | 38.2 (± 6.12) | 10.44 (± 3.54) | 14.28 (± 3.47) |
| **LRRK2 DM** | 30.68 (± 4.04) | 35.58 (± 6.52) | 8.96 (± 2.56) | 13.02 (± 3.40) |

**Fig. S5. Representative confocal image of colocalization of ectopically expressed NOD2 with LRRK2.** Colocalization of LRRK2 wild-type (WT) and CRISPR/Cas-edited LRRK2 double-mutant (DM, N551K+R1398H) in cells transfected with plasmids expressing NOD2 WT (**A**) and NOD2 R702W (**B**). (**C**) LRRK2 KO cells (top panel) and untransfected LRRK2 WT cells (bottom panel) were used as a negative control for antibody specificity. (**A**-**D**) RAW264.7 cell lines with the three LRRK2 genotypes were transfected with NOD2 plasmid and a LRRK2 WT cell line was kept untransfected. Twenty-four hours after electroporation, the transfected and untrasfected cells were treated with or without 10 μg/mL of N-glycolyl MDP for another 24 hours.  Cells were fixed with 4% paraformaldehyde, permeabilized and double stained for LRRK2 and NOD2 with rabbit anti-LRRK2 (1:500) and mouse anti-FLAG (1:250) antibodies. Nuclei were stained with DAPI. Images were obtained by confocal microscopy.

Colocalization between LRRK2 and NOD2 was measured from 25-30 cells by Zeiss 2012 ZEN confocal software (**D**).

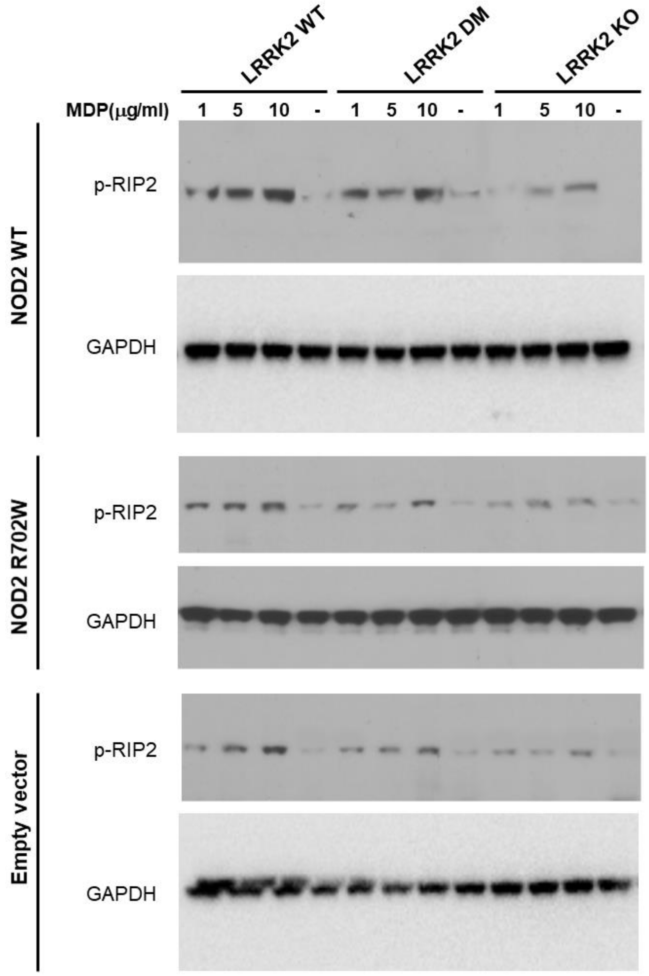


**Fig. S6. Effects of LRRK2 variants on MDP-induced phosphorylation of RIP2 at Ser 176.** LRRK2 Wild-type (WT), CRISPR/Cas-edited LRRK2 double-mutant (DM, N551K+R1398H) and LRRK2 knock-out (KO) RAW264.7 cell lines were transfected with plasmids expressing NOD2 WT, NOD2 mutant (R702W) or an empty vector [pcDNA3.1+/C-(K)DYK] as a control. Twenty-four hours post-transfection, cells were left untreated or treated with different concentrations of N-glycolyl MDP as indicated, for another 24 hours. Cell lysates were prepared and the phosphorylation of RIP2 (p-RIP2) in the transfected cell lines was analyzed by immunoblotting with a specific antibody against RIP2 when phosphorylated at Ser 176. GAPDH was used as a loading control.

#### Supplementary Tables

**Table S1.** Summary of whole genome sequencing data and mapping quality control of six samples from the studied family.

| Parameters | ID2 | ID3 | ID4 | ID5 | ID6 | ID7 | Average |
|---|---|---|---|---|---|---|---|
| No. paired-end reads (millions) | 471 | 416 | 350 | 520 | 437 | 526 | **453** |
| Mapped reads | 99.1% | 98.9% | 99.2% | 99.4% | 99.1% | 99.4% | **99.2%** |
| Mean Mapping Quality | 51.0 | 51.0 | 51.2 | 51.1 | 51.0 | 51.0 | **51.0** |
| Overlapping read pairs | 4.06% | 5.84% | 5.08% | 3.78% | 4.05% | 3.80% | **4.4%** |
| Duplicated reads | 12.2% | 12.2% | 11.1% | 12.1% | 10.8% | 11.3% | **11.6%** |
| Coverage (mean X ± SD)* | 32 ± 12 | 28 ± 11 | 25 ± 10 | 35 ± 13 | 30 ± 12 | 36 ± 13 | **31 ± 12** |
| Genome fraction with ≥ 10X* | 91.4% | 91.3% | 91.5% | 91.4% | 91.3% | 91.4% | **91.4%** |
| Genome fraction with ≥ 20X* | 90.3% | 89.3% | 82.0% | 90.7% | 90.0% | 90.8% | **88.8%** |
| Genome fraction with ≥ 30X* | 72.0% | 56.7% | 32.1% | 83.5% | 68.1% | 85.3% | **66.3%** |

* Overlapping read pairs and duplicated reads were ignored.
No.: number; SD: standard deviation; X: times/folds.

**Table S2**. Candidate single nucleotide variants (SNVs) and short Indels identified in the studied family by applying the custom filtering approaches shown in Figure S1.

| Gene | GDI [a] | Chr | Position [b] | Ref | Alt | Type | rsID [a] | MAF (ExAC ALL) | AA Change | Scaled CADD [a] | PolyPhen-2 HumVar [a] | Prioritized [c] |
|---|---|---|---|---|---|---|---|---|---|---|---|---|
| **Approach #1: Recessive model - All cases in the family, regardless of age-at-diagnosis (ID2, ID4, ID6 and ID7 are homozygous for the variant)¶.** | | | | | | | | | | | | |
| *HLA-A* | High | 6 | 29913037 [d] | G | A | Missense | rs1137631 | 15.69% | V358M | 15.43 | Benign | No |
| **Approach #2: Recessive model - Younger cases in the family (ID4, ID6 and ID7 are homozygous for the variant) ¶.** | | | | | | | | | | | | |
| *HLA-A* | High | 6 | 29910719 | G | A | Missense | rs2230991 | 15.65% | E87K | 9.87 | Benign | No |
| *HLA-A* | High | 6 | 29910721 | G | C | Missense | rs199474424 | 15.56% | E87D | 14.42 | Benign | No |
| *RNF39* | Medium | 6 | 30039418 | C | T | Missense | rs2301752 | 12.27% | A245T | 7.02 | Benign | No |
| *KIAA1217* | Medium | 10 | 24813454 | G | A | Missense | rs10828663 | 22.63% | A887T | 14.18 | Benign | No |
| *RNH1* | Medium | 11 | 499120 [d] | G | A | Missense | rs17585 | 11.39% | P170L | 16.60 | Benign | No |
| *LRRK2* | Medium | 12 | 40657700 [d] | C | G | Missense | rs7308720 | 8.61% | N551K | 24.1 | Probably damaging | Yes |
| *PLD2* | High | 17 | 4722876 | G | A | Missense | rs3764897 | 15.98% | G821S | 21.30 | Benign | No |
| *CD68* | Medium | 17 | 7484101 [d] | C | A | Missense | rs9901673 | 15.45% | Q254K | 12.1 | Benign | No |
| *MPDU1* | Medium | 17 | 7490810 [d] | G | A | Missense | rs10852891 | 15.40% | A229T | 20.6 | Benign | No |
| *SALL4* | Medium | 20 | 50406630 [d] | T | G | Missense | rs6091375 | 5.10% | I798L | 7.489 | Benign | No |
| *TLR7* | Medium | X | 12903659 [d] | A | T | Missense | rs179008 | 18.01% | Q11L | 5.466 | Benign | No |
| **Approach #3: Recessive model - Early-onset cases in the family (Only ID6 and ID7 are homozygous for the variant) ¶.** | | | | | | | | | | | | |
| *ZNF678* | Medium | 1 | 227843003 | G | A | Missense | rs61740826 | 2.09% | C406Y | 22.4 | Probably damaging | Yes |
| *LRP1B* | Medium | 2 | 141242918 [d] | T | C | Missense | rs34488772 | 4.78% | Q3140R | 0.121 | Benign | No |
| *PRIMPOL* | Medium | 4 | 185580557 [d] | A | G | Missense | rs74696256 | 0.90% | T82A | 22.5 | Benign | No |
| *ROS1* | High | 6 | 117710661 [d] | T | C | Missense | rs28639589 | 4.59% | I537M | 17.8 | Benign | No |
| *LRRK2* | Medium | 12 | 40702911 [d] | G | A | Missense | rs7133914 | 8.41% | R1398H | 23.2 | Possibly damaging | Yes |
| *STAB2* | Medium | 12 | 103988285 [d] | A | G | Missense | rs17034186 | 1.16% | I110V | 17.34 | Benign | No |
| *SOS2* | Medium | 14 | 50655307 [d] | C | T | Missense | rs61755579 | 1.85% | A208T | 23 | Benign | No |
| *SMPD3* | Medium | 16 | 68395522 [d] | C | T | Missense | rs71395853 | 3.87% | C617Y | 18.13 | Benign | No |
| *DHX33* | Medium | 17 | 5354204 [d] | G | C | Missense | rs11653658 | 5.59% | H483D | 17.74 | Benign | No |
| *HRH4* | Medium | 18 | 22057204 [d] | C | G | Missense | rs58154316 | 1.08% | S284C | 16 | Probably damaging | No |
| *ACP5* | Medium | 19 | 11687195 | C | T | Missense | rs2229531 | 9.66% | V200M | 22.0 | Possibly damaging | Yes |
| *ACP5* | Medium | 19 | 11687351 [d] | C | T | Missense | rs2305799 | 10.34% | V148M | 15.69 | Benign | No |
| *LYL1* | Medium | 19 | 13211843 | C | T | Missense | rs117072928 | 8.93% | R48Q | 1.86 | Benign | No |
| *KLK8* | Medium | 19 | 51503285 [d] | C | T | Missense | rs16988799 | 4.64% | V154I | 13.99 | Benign | No |
| *DIDO1* | Medium | 20 | 61512185 | G | C | Missense | rs41282984 | 6.40% | S1708C | 10.96 | Benign | No |
| *SLC17A9* | Medium | 20 | 61598731 | C | T | Missense | rs7271712 | 2.65% | T397M | 24.2 | Possibly damaging | Yes |
| *PRDM15* | Medium | 21 | 43221797 [d] | G | C | Missense | rs2236695 | 3.83% | T1376S | 16.96 | Benign | No |
| *TCF20* | Medium | 22 | 42607817 [d] | C | T | Missense | rs17002890 | 0.99% | M1165I | 22.8 | Benign | No |
| **Approach #4: Recessive model - Early-onset cases in the family (ID6 and ID7 are compound heterozygous).** | | | | | | | | | | | | |
| *NPHP4* | Medium | 1 | 5937289 | T | C | Missense | rs113097479 | 0.13% | H894R | 0.001 | Benign | No |
| | | | 5965440 | T | C | Missense | rs35959882 | 0.04% | T623A | 0.001 | Benign | |
| *C1orf167* | - | 1 | 11826186 | G | T | Missense | rs145919329 | 0.18% | A115S | 13.2 | - | No |
| | | | 11826351 | G | A | Missense | rs144306270 | 0.20% | G170R | 2.912 | - | |
| | | | 11826421 | A | G | Missense | rs187345781 | 0.17% | H193R | 0.001 | - | |
| | | | 11826861 | A | C | Missense | rs188115585 | - | N340H | 4.995 | - | |
| | | | 11827071 | C | T | Missense | rs189594838 | 0.78% | R410W | 13.49 | - | |
| | | | 11827114 | AC | - | Frameshift deletion | rs141465134 | 0.28% | D424fs | 16.87 | - | |
| | | | 11847932 | T | C | Missense | rs116698217 | 0.23% | C1226R | 2.025 | Benign | |
| *IQGAP3* | High | 1 | 156518421 [d] | C | T | Missense | rs77834544 | 0.21% | G649S | 0.317 | Benign | No |
| | | | 156521798 [d] | T | A | Missense | - | - | Q513L | 17.8 | Benign | |
| *CCDC141* | Medium | 2 | 179702426 [d] | C | G | Missense | rs75153675 | 0.007% | E1174Q | 16.69 | Benign | No |
| | | | 179839888 [d] | G | A | Missense | rs10497529 | 0.0259 | A141V | 27.3 | Probably damaging | |
| *INTU* | Medium | 4 | 128608951 [d] | G | A | Missense | - | 0.001% | D460N | 22.2 | Benign | No |
| | | | 128627927 [d] | T | G | Missense | rs34311863 | 0.31% | C692G | 15.57 | Benign | |
| *SORBS2* | Medium | 4 | 186545346 [d] | A | T | Missense | rs61736043 | 0.36% | L509I | 0.937 | Probably damaging | No |
| | | | 186599973 [d] | C | T | Missense | rs190199282 | 0.04% | R36H | 22.1 | Probably damaging | |
| *FAT1* | High | 4 | 187524714 [d] | C | T | Missense | rs192691397 | 0.05% | V3656I | 16.21 | Benign | No |
| | | | 187530423 [d] | T | C | Missense | rs138364727 | 0.007% | I3374V | 23.9 | Probably damaging | |
| | | | 187540374 [d] | C | T | Missense | rs370340394 | 0.001% | A2456T | 6.944 | Benign | |
| | | | 187628947 [d] | C | T | Missense | rs61733571 | 0.27% | V679I | 26.3 | Probably damaging | |
| *CDC20B* | Medium | 5 | 54468432 [d] | CTTCT | - | Frameshift deletion | rs137940833 | 0.31% | R36fs | 26.1 | - | No |
| | | | 54468450 [d] | T | C | Missense | rs138811807 | 1.15% | D31G | 11.88 | Benign | |
| *APC* | Medium | 5 | 112043492 | C | A | Missense | rs113782655 | 0.15% | S26R | 19.74 | Benign | No |
| | | | 112102943 | T | A | Missense | - | - | L93H | 24.1 | Possibly damaging | |
| | | | 112102945 | C | G | Missense | - | - | R94G | 23.1 | Benign | |
| *DMXL1* | Medium | 5 | 118469561 [d] | G | A | Missense | rs139365266 | 0.20% | V648I | 21.3 | Benign | No |
| | | | 118485204 [d] | G | A | Missense | rs140855219 | 0.03% | V1228M | 28 | Probably damaging | |
| *TNXB* | High | 6 | 32029369 | C | T | Missense | rs200135227 | 0.25% | V2433I | 10.89 | Benign | No |
| | | | 32049373 | C | T | Missense | - | 0.005% | V1272M | 14.38 | Benign | |
| | | | 32063558 | C | T | Missense | rs201146825 | 0.007% | G691D | 23.7 | Probably damaging | |
| *PKHD1* | Medium | 6 | 51712759 [d] | T | C | Missense | rs7766366 | 0.25% | T2641A | 22.7 | Probably damaging | No |
| | | | 51875133 [d] | G | A | Missense | rs115338476 | 0.18% | R1909W | 21.2 | Benign | |
| | | | 51890265 [d] | T | C | Missense | rs116809571 | 0.20% | E1448G | 12.2 | Benign | |

| | | | | | | | | | | | | |
|---|---|---|---|---|---|---|---|---|---|---|---|---|
| | | | 51917987 [d] | G | C | Missense | rs115045643 | 0.22% | P676R | 0.044 | Benign | |
| *TINAG* | Medium | 6 | 54173421 [d] | T | G | Missense | rs34700914 | 0.26% | S25A | 1.127 | Benign | No |
| | | | 54214618 [d] | C | T | Missense | rs139989527 | 0.01% | T335M | 28.6 | Probably damaging | |
| *EYS* | High | 6 | 65300527 | C | T | Missense | rs145274061 | 0.02% | D1745N | 8.095 | Benign | No |
| | | | 66005791 | C | T | Missense | - | - | G663E | 14.67 | Benign | |
| *SGK223* | High | 8 | 8176654 | G | C | Missense | rs2011560 | 0.24% | H1077Q | 0.056 | Benign | No |
| | | | 8235555 | G | T | Missense | rs55764617 | 0.33% | L122I | 11.19 | Benign | |
| *DDX31* | Medium | 9 | 135470281 | C | T | Missense | rs306548 | 0.35% | R843Q | 7.707 | Benign | No |
| | | | 135538016 | C | T | Missense | rs17402080 | 1.18% | E153K | 0.363 | Benign | |
| *PDCD11* | High | 10 | 105183348 [d] | T | C | Missense | rs61751511 | 0.87% | V899A | 25.5 | Possibly damaging | No |
| | | | 105201712 [d] | G | A | Missense | - | - | E1563K | 23 | Benign | |
| *SLC22A24* | Medium | 11 | 62886706 [d] | C | T | Missense | rs116063135 | 0.41% | R203H | 22.2 | Probably damaging | Yes |
| | | | 62910891 [d] | C | A | Missense | rs116409312 | 0.31% | V121L | 23 | Probably damaging | |
| *GCN1* | - | 12 | 120574343 [d] | G | A | Missense | rs201840533 | 0.02% | A2324V | 21.1 | Benign | No |
| | | | 120574344 [d] | C | A | Missense | - | - | A2324S | 22.9 | Benign | |
| | | | 120613593 [d] | G | A | Missense | rs114251901 | 0.14% | S333L | 18.24 | Benign | |
| *KL* | Medium | 13 | 33590898 | C | T | Missense | rs115511178 | 0.19% | A107V | 12.57 | Benign | No |
| | | | 33590971 | C | A | Missense | rs79554512 | 0.13% | N131K | 18.81 | Possibly damaging | |
| *AHNAK2* | High | 14 | 105414032 [d] | G | T | Missense | rs199905726 | 0.13% | L2586I | 0.075 | Benign | No |
| | | | 105414053 [d] | T | C | Missense | rs200965573 | 0.11% | M2579V | 1.787 | Benign | |
| | | | 105419551 [d] | C | G | Missense | rs201524595 | 2.49% | G746A | 11.17 | Possibly damaging | |
| *ABCC6* | Medium | 16 | 16259497 [d] | G | T | Missense | rs60707953 | 0.31% | L1097I | 16.29 | Possibly damaging | No |
| | | | 16259722 [d] | G | C | Missense | rs57179857 | 0.07% | Q1022E | 17.15 | Benign | |
| *ZNF469* | Medium | 16 | 88496285 | G | T | Missense | rs113484918 | 0.35% | A803S | 1.577 | Benign | No |
| | | | 88496544 | C | T | Missense | rs145186655 | 0.22% | A889V | 6.999 | Benign | |
| | | | 88504208 | G | C | Missense | rs569602115 | 0.03% | G3416R | 15.84 | Possibly damaging | |
| | | | 88504673 | G | T | Missense | rs199760004 | 0.14% | A3571S | 5.491 | Benign | |
| *ATAD5* | High | 17 | 29162972 | G | C | Missense | rs144812489 | 0.01% | A625P | 16.31 | Possibly damaging | No |
| | | | 29187582 | C | T | Missense | rs35910070 | 0.21% | L1030F | 17.44 | Possibly damaging | |
| *GPR179* | Medium | 17 | 36485883 | C | T | Missense | rs80172972 | 0.33% | R1190Q | 0.481 | Benign | No |
| | | | 36487060 | C | T | Missense | rs78470373 | 0.38% | A798T | 19.39 | Benign | |
| **Approach #5: Dominant model - All cases in the family, regardless of age-at-diagnosis (ID2, ID4, ID6 and ID7 are heterozygous for the variant).** | | | | | | | | | | | | |
| *PPP4R2* | Medium | 3 | 73114043 [d] | C | T | Missense | - | - | P227S | 17.9 | Benign | No |
| *CP** | Medium | 3 | 148905977 [d] | T | C | Missense | - | - | K576E | 26.6 | Possibly damaging | Yes |
| *SOWAHB* | Medium | 4 | 77816977 [d] | G | C | Missense | - | - | L676V | 24.6 | Possibly damaging | Yes |
| *SYNPO2** | High | 4 | 119947864 [d] | G | A | Missense | - | - | E114K | 22.1 | Benign | No |
| *UGT3A1* | Medium | 5 | 35957503 [d] | C | T | Missense | - | - | A288T | 8.7 | Benign | No |
| *HLA-DRB1* | High | 6 | 32552091 [d] | G | C | Missense | rs569286159 | 2.67% | F55L | 1.916 | Benign | No |
| *MCM9** | Medium | 6 | 119147418 [d] | G | A | Missense | - | - | T618I | 25.6 | Possibly damaging | Yes |
| *CCDC34* | Medium | 11 | 27384674 | C | A | Missense | - | - | R23I | 18.45 | Benign | No |
| *LEMD3* | Medium | 12 | 65563747 | G | A | Missense | - | - | G124D | 16.9 | Benign | No |
| *ERN1* | Medium | 17 | 62130282 [d] | T | C | Missense | - | - | K704R | 12.88 | Benign | No |
| *CDH20** | Medium | 18 | 59217360 [d] | G | A | Missense | - | - | D600N | 8.2 | Benign | No |
| *SUV39H1* | Medium | X | 48564979 | C | T | Missense | - | - | R356W | 17.12 | Benign | No |
| *ZNF185* | Medium | X | 152113882 [d] | C | G | Missense | - | - | A459G | 10.8 | Benign | No |
| **Approach #6: Dominant model - Younger cases in the family (ID4, ID6 and ID7 are heterozygous for the variant).** | | | | | | | | | | | | |
| *PRKACB* | Medium | 1 | 84650810 [d] | G | T | Stop-gain | - | - | E122X | 37 | - | Yes |
| *IQGAP3* | High | 1 | 156521798 [d] | T | A | Missense | - | - | Q513L | 17.8 | Benign | No |
| *ASTN1* | Medium | 1 | 176833514 [d] | T | G | Missense | - | - | D1264A | 20.1 | Benign | No |
| *CR1* | Medium | 1 | 207785097 [d] | A | G | Missense | - | - | E1674G | 21.8 | Possibly damaging | Yes |
| *FSIP2* | High | 2 | 186666910 [d] | C | G | Missense | - | - | H4382D | 15 | - | No |
| *NBEAL2* | High | 3 | 47037443 [d] | G | A | Missense | - | - | V685M | 22.0 | Possibly damaging | No |
| *OTOP1* | Medium | 4 | 4199651 [d] | T | A | Missense | - | - | M304L | 0.174 | Benign | No |
| *SLC25A25* | Medium | 9 | 130830773 | G | A | Missense | - | - | V59I | 21.1 | Benign | No |
| *PDCD11* | High | 10 | 105201712 [d] | G | A | Missense | - | - | E1563K | 23 | Benign | No |
| *ADAL* | Medium | 15 | 43627339 [d] | A | G | Missense | - | - | T12A | 16.0 | Benign | No |
| *CCPG1* | Medium | 15 | 55670559 [d] | C | A | Missense | - | - | G64V | 9.7 | Benign | No |
| *TRIP4* | Medium | 15 | 64737241 [d] | G | A | Missense | - | - | V538I | 23.2 | Benign | No |
| *RBBP6* | Medium | 16 | 24583228 [d] | C | T | Missense | - | - | P1614L | 23.6 | Benign | No |
| *NOD2* | Medium | 16 | 50745926 [d] | C | T | Missense | rs2066844 | 2.27% | R702W | 24.6 | Possibly damaging | Yes |
| *RBL2* | Medium | 16 | 53515590 [d] | C | T | Missense | - | - | A1031V | 19.3 | Benign | No |
| *LRRC25* | Medium | 19 | 18507073 [d] | G | A | Missense | - | - | P234L | 1.9 | Benign | No |
| *BCKDHA* | - | 19 | 41916587 [d] | C | T | Missense | - | - | P52S | 23 | Benign | No |
| *SCAF1* | Medium | 19 | 50156323 | G | A | Missense | - | - | G893S | 22.9 | Benign | No |
| *SCAF1* | Medium | 19 | 50156330 | C | A | Missense | - | - | T895N | 12.8 | Benign | No |
| *ISOC2* | Medium | 19 | 55964727 [d] | G | - | Frameshift deletion | - | - | P119fs | 34 | - | Yes |
| *ZIM2* | Medium | 19 | 57301244 [d] | G | A | Missense | - | - | S158F | 6.164 | Benign | No |
| *NSFL1C* | Medium | 20 | 1434931 [d] | T | A | Missense | - | - | Y155F | 28.3 | Probably damaging | Yes |
| *CRELD2* | Medium | 22 | 50315340 [d] | C | T | Missense | - | - | P175S | 0.203 | Benign | No |
| **Approach #7: Dominant model - Early-onset cases in the family (*de novo* variant: only ID6 and ID7are heterozygous for the variant).** | | | | | | | | | | | | |
| *NUP153* | High | 6 | 17625023 [d] | C | A | Missense | - | - | A1315S | 23.9 | Possibly damaging | No |

AA: amino acid; Alt: Alternative allele; CADD: Combined annotation dependent depletion; Chr: Chromosome, ExAC: Exome Aggregation Consortium; GDI: Gene damage index; MAF: Minor allele frequency; Ref: Reference allele.

**a** dbSNP (2016), GDI (January 2016), CADD v1.4 and PolyPhen-2 v2.2.2r398.

**b** Genomic position on GRCh37.

**c** Candidate variants were prioritized if they presented all the three following criteria: i) the variant had CADD $\geq$ 20, ii) the variant was a predicted damaging missense variant (PolyPhen-2), nonsense variant or frameshift indel and iii) the variant was located in a gene with low/medium GDI score. In approach #4, variants were prioritized if the two or more compound heterozygous variants reached criteria i and ii, while the gene reached criteria iii. Prioritized variants are highlighted in yellow in the table.
**d** Genotypes from WGS were validated by WES.
¶ Chr X: male carries the variant and female is homozygous for the variant.

* Subject ID3 also carries the same mutation in this gene as the affected members of the family.

**Table S3.** Candidate deletion structural variants (DSVs) identified in the studied family by applying the custom filtering approaches shown in Figure S1.

| Chr | Start** | End** | Length (Kb) | Gene | Gene overlap | SV overlap# | Observed loss¶ |
|---|---|---|---|---|---|---|---|
| **Approach #5: Dominant model - All cases in the family, regardless of age-at-diagnosis (ID2, ID4, ID6 and ID7 are heterozygous for the variant).** | | | | | | | |
| **5** | 132918958 | 132925014 | 6.06 | *FSTL4* | CDS | esv2663891 | 398/1151 |
| **18** | 46997685 | 47005333 | 7.65 | *C18orf32* | EXON | esv3642494 | 203/2504 |
| **22** | 44564926 | 44566040 | 1.11 | *PARVB* | EXON | esv3647889 | 213/2504 |
| **Approach #6: Dominant model - Younger cases in the family (ID4, ID6 and ID7 are heterozygous for the variant).** | | | | | | | |
| **9** | 132373830 | 132374787 | 0.96 | *C9orf50* | CDS | esv3621837 | 14/2504 |

**Genomic position on GRCh37.

### Accession number of structural variant detected in The 1000 genomes consortium that overlaps with the DSV in this study.

¶ Number of samples with SV (homozygous + heterozygous)/ Total number of samples sequenced by The 1000 genomes consortium.

**Table S4.** Oligonucleotides used in the CRISPR/Cas construct of *Lrrk2* variants.

| **Forward and reverse target DNA oligonucleotides** | **Sequences** |
|---|---|
| N551K forward | 5’-TAATACGACTCACTATAGGAACAGGGTATGTAGA-3’ |
| N551K reverse | 5’-TTCTAGCTCTAAAACTTGTTCTACATACCCTGTT-3’ |
| R1398H forward | 5’-TAATACGACTCACTATAGAGAATTCCTCACGACC-3’ |
| R1398H reverse | 5’-TTCTAGCTCTAAAACTCTAGGTCGTGAGGAATTC-3’ |
| **Donor HDR templates** | **Sequences** |
| N551K | 5’A*G*T*AAACAACAAGAAAGTAAAGTAAAACTCAAAGCCCCA CCCCCAGACCCTCAGATGTTAGTCTCGTTAATGGTATAAAGA CAGAAAATCCTTGTTCTACATACCCTCTTCAGAGCGACTAGA ACCAGCTTGTGGATGTCAGTCCTGAAACACTGTTTTCTGAGC ACATTTGGTCTGCATTGAGA*G*T*C-3’ |
| R1398H | 5’-A*G*A*TCATAGACAGCCAGGTAGAGGGCTCTCTGGGTCAT GAAGTGGGGGTGAGTGCTGTAGAATTCCTCACGACCTAGA AGGAGATATCAGAGGTTTGAGTCTTTCCCATAGTAGGTAGG ACTCGTTACGAAATAAT*G*A*G-3’ |
| **Sequences of PCR primers** | **Sequences** |
| N551K forward | 5’-GGCAGTGTGTGGAGCCTAAA-3’ |
| N551K reverse | 5’-GGCATCAGAGAAGACAGCCA-3’ |
| R1398H forward | 5’-GTCCATCCAAATACGGGGCA-3’ |
| R1398H reverse | 5’-GGGCATCCAGGGACACATAA-3’ |

*Phosphorothioate modification
